# Understanding the contexts and mechanisms of bluespace prescription programmes implemented in health and social care settings: a realist review

**DOI:** 10.1101/2021.10.15.21264908

**Authors:** Julius Cesar Alejandre, Sebastien Chastin, Katherine N. Irvine, Michail Georgiou, Preeti Khanna, Zoë Tieges, Niamh Smith, Yong-Yee Chong, Frances Claire Onagan, Lesley Price, Sharon Pfleger, Rachel Helliwell, Judith Singleton, Samuel Curran, Karin Helwig

**Affiliations:** Department of Civil Engineering and Environmental Management, School of Computing, Engineering and Built Environment, Glasgow Caledonian University, Glasgow, United Kingdom; Department of Physiotherapy and Paramedicine, School of Health and Life Sciences, Glasgow Caledonian University, Glasgow, United Kingdom; Department of Movement and Sport Sciences, Ghent University, Ghent, Belgium; Social, Economic and Geographical Sciences Department, The James Hutton Institute, Aberdeen, United Kingdom; Department of Food and Nutrition, Institute of Home Economics, University of Delhi, India; SMART Technology Centre, School of Computing, Engineering and Built Environment, Glasgow Caledonian University, Glasgow, Scotland, United Kingdom; Geriatric Medicine, Usher Institute, University of Edinburgh, Edinburgh, Scotland, United Kingdom; Centre for Sport and Exercise Sciences, University of Malaya, Kuala Lumpur, Malaysia; Department of Health, Health Promotion Bureau, Manila, Philippines; Department of Nursing and Community Health, School of Health and Life Sciences, Glasgow Caledonian University, Glasgow, United Kingdom; Public Health Directorate, National Health Service Highland, Inverness, United Kingdom; School of Pharmacy, The Robert Gordon University, Aberdeen, United Kingdom; Centre of Expertise for Waters, The James Hutton Institute, Aberdeen, United Kingdom; Faculty of Health, School of Clinical Sciences, Queensland University of Technology, Brisbane, Queensland, Australia; International Services, Scottish Environment Protection Agency, Stirling, United Kingdom

**Keywords:** sustainable healthcare, social prescribing, blue spaces, bluespace prescriptions

## Abstract

**Background:** Nature-based social prescribing programmes such as ‘bluespace prescription’ may promote public health and health improvement of individuals with long-term conditions. However, there is limited systematically synthesised evidence that investigates the contexts and mechanisms of Bluespace Prescription Programmes (BPPs) that could inform programme theories for policy and practice.

**Methods:** We conducted a realist review by searching six databases for articles published between January 2000 and February 2020, in English, about health and social care professionals providing referral to or prescription of blue space activities with health-related outcomes. We developed themes of contextual factors by analysing the contexts of BPPs. We used these contextual factors to develop programme theories describing the mechanisms of BPP implementation. Our study was registered with PROSPERO (CRD42020170660).

**Results:** Fifteen studies with adequate to strong quality were included from 6,736 records. Service users had improvements on their physical, mental, social health, and environmental knowledge after participating in BPPs referred to or prescribed by health and social care professionals. Patient-related contextual factors were referral information, free equipment and transportation, social support, blue space environments, and skills of service providers. Programme-related contextual factors were communication, multi-stakeholder collaboration, financing, and adequate service providers. Programme theories on patient enrolment, engagement, adherence, communication protocols, and long-term programme sustainability described the mechanisms of BPP implementation.

**Conclusion:** BPPs could support health and social care services if contextual factors and mechanisms of programme theories associated with patients and programme delivery are considered for implementation. Our findings have implications in planning, development, and implementation of similar nature-based social prescribing programmes in health and social care settings.

## Introduction

Social prescribing (SP) is a health and social care approach that promotes health by connecting individuals to community-based and non-medical programmes providing physical activity and social support (Teuton, 2015; Husk et al., 2020). SP is delivered through four referral pathways: 1) signposting; 2) direct referral; 3) link worker, and 4) holistic link worker (Husk et al., 2020). The process commonly starts with a consultation with a healthcare worker (general practitioner/GP). If GPs give information through leaflets and direct patients to community-based organisations (CBOs) or service providers, this is referred to as signposting. Direct referral is when GPs contact CBOs and arrange patient enrolment. The link worker pathway comprises GPs connecting patients to link workers who arrange patient enrolment. The holistic link worker pathway includes a feedback loop between CBOs and link workers (Husk et al., 2020). Moreover, SP implementation is described by programme theories (Husk et al., 2020; Pawson et al., 2005; Rycroft-Malone et al., 2012) on patient enrolment (first successful referral), engagement (attendance to first session), adherence (maintained participation over time), link worker coordination (communication), and partnerships with CBOs (supportive interventions) (Husk et al., 2020; Bickerdike et al., 2017; Chatterjee et al., 2018; Bertotti et al., 2018). SP could help motivate individuals to engage in health-promoting behaviours such as physical activity, socialisation, and self-management (Husk et al., 2020; Bickerdike et al., 2017; Bertotti et al., 2018). It could also help reduce primary healthcare burden by depressurising healthcare professionals’ workload through promotion of non-clinical interventions (Bertotti et al., 2018).

Nature-based social prescribing or nature prescription is a type of SP that promotes health and wellbeing through contact with nature (Husk et al., 2020; Leavell et al., 2019). Exposure to nature can improve physical and mental health (Hartig et al., 2014), and the presence of blue spaces, defined as natural and manmade outdoor surface water environments, enhances these effects (Pouso et al., 2021; Benton et al., 2021; Völker and Kistemann, 2011). Contact with blue spaces are associated with health outcomes as illustrated by the ‘Blue Space and Health/Wellbeing Model’ (White et al., 2020). Exposure to blue spaces in terms of duration, frequency, and intensity is associated with individuals’ contact with blue spaces which could be indirect (window view), incidental (commute along a river path), or intentional (beach visit) (White et al., 2020; Keniger et al., 2013). Contact with blue spaces improves health outcomes through mitigation (noise abatement), instoration (physical activity), and restoration (stress reduction) pathways (White et al., 2020; Markevych et al., 2017), resulting in improved mood, self-esteem, physical activity, social interactions, and decrease in mortality (Georgiou et al., 2021; Tieges et al., 2020; Finlay et al., 2015). These pathways are associated with environmental (type, quality, weather) or personal (age, gender, ethnicity) effect modifiers influencing engagement with blue space activities (BSAs) (White et al., 2020). The exposure– outcome pathway is catalysed by feedback actions such as quality improvement (societal) and safe access (local) to blue spaces (White et al., 2020). It also includes personal actions such as nature-based social prescribing (Blue Gyms and prescribing therapeutic BSAs) that targets individuals’ motivations to have contact with blue spaces (White et al., 2020; Depledge and Bird, 2009; Britton et al., 2020).

The knowledge base on social prescribing falls behind the fast growth and roll-out of social prescribing programmes on the ground (Husk et al., 2020). There is limited understanding on the contextual implementation of specific types of SP (Husk et al., 2020) including nature-based social prescribing programmes the utilise blue spaces. There is also an evidence gap on the effectiveness (Bickerdike et al., 2017; Chatterjee et al., 2018; Pescheny et al., 2018), suitability (Pilkington et al., 2017), referral processes (Chatterjee et al., 2018), and factors influencing SP uptake (Britton et al., 2020; Buckley and Brough, 2017). Additionally, existing systematic reviews on social prescribing mainly focused on general and greenspace programmes (Husk et al., 2020; Bertotti et al., 2018; Masterton et al., 2020). Considering these gaps and the growing evidence on the health benefits of blue spaces, we conducted a realist review to investigate the contexts and mechanisms of implementing Bluespace Prescription Programmes (BPPs) delivered to people with health conditions. We used realist synthesis (Pawson et al., 2005) to investigate the contextual factors influencing BPP implementation (Pawson et al., 2005; Rycroft-Malone et al., 2012) and used these contextual factors to develop programme theories describing the mechanisms of BPP implementation.

## Methods

Our systematic realist review followed the PRISMA guidelines (Page et al., 2021) and realist reviews (Berg, 2016). A realist review is way of evidence synthesis that provides explanation why interventions work or do not work in certain circumstances by interrogating the contextual complexities and mechanisms of implementing such interventions (Rycroft-Malone, 2012). The steps of a realist review are: 1) identifying the question; 2) clarifying the purpose of the review; 3) finding and articulating the initial programme theories; 4) searching for evidence; 5) appraising the evidence; 6) extracting the data; 7) synthesising findings; and 8) drawing conclusions and recommendations (Berg, 2016).

### Identifying the question and clarifying the purpose of the review

We defined BPP as individual or group activities that take place in or around natural water environments such as surfing, swimming, running near blue spaces, amongst others (interventions), which are referred to or prescribed by health and social care professionals (population or participants) through SP referral pathways and with health-related outcomes (outcomes) (Husk et al., 2020). We included peer-reviewed research on BPPs conducted in natural water environments which were referred to or prescribed by health and social care workers (GPs, pharmacists, social workers, link workers, etc.) to individuals attending health and social care facilities and with health-related outcomes. We included case reports, qualitative, case-control, cohort, pre-post intervention studies, non-randomised, and randomised controlled trials published in English. We limited the publication period from January 2000 to February 2020 since research on the health benefits of blue spaces emerged in the early 2000s (Völker and Kistemann, 2011; Britton et al., 2020). We excluded studies where non-health or non-social care workers referred BSAs to participants through advertisements and studies in which BSAs were not conducted in natural water environments.

### Finding and articulating the initial programme theories

Our search and data analysis were guided by the following initial social prescribing programme theories (PTs):

Initial PT1: If referral is presented in an acceptable manner, it is compatible with the patient’s needs and expectations, and the patient believes that it will improve their condition, then they may enrol (enrolment) (Husk et al., 2020).

Initial PT2: If transportation is provided making the socially prescribed activity accessible to the patient, then they will engage (engagement) (Husk et al., 2020).

Initial PT3: If the service providers are skilled and there are improvements on patient’s condition, then they are more likely to keep attending (adherence) (Husk et al., 2020).

Initial PT4: If there are link workers facilitating delivery of social prescriptions and liaising with health and social care facilities and third sector organisations, then the referral uptake will be successful (link worker coordination) (Husk et al., 2020; Bickerdike et al., 2017; Chatterjee et al., 2018; Bertotti et al., 2018).

Initial PT5: If partnership between health and social care facilities and community-based organisations is financially supported, then the delivery of social prescribing programmes will be sustained (partnership with CBOs) (Bickerdike et al., 2017; Chatterjee et al., 2018; Bertotti et al., 2018).

### Searching for evidence

We searched PubMed, Web of Science, PsycInfo, MEDLINE, Scopus, and CINAHL using keyword strings (Appendix 1) and conducted snowball search by screening the reference lists of included studies (Schardt et al., 2007; Jalali and Wohlin, 2012). Records were uploaded, de-duplicated, and screened using Rayyan QCRI (Ouzzani et al., 2016). Title, abstract (JA, KH, SCh, and 17 researchers), and full-text screenings (JA–KH, JA–SCh) were independently conducted in pairs. KNI resolved conflicting decisions.

### Appraising the evidence

Included studies were categorised based on their methodological approaches to suit the QualSyst tools (Kmet et al., 2004). Quality of studies was independently assessed in pairs (JA– KH, JA–SCh, JA–KNI). Conflicting ratings were reconciled by one-to-one discussion. We adopted strong (≥0·80), good (0·71–0·79), adequate (0·51–0·70), and limited (≤0·50) quality thresholds for quantitative papers (Lee et al., 2008) and used adequate (≥0·55) and low (≤0·54) for qualitative studies (Maharaj and Harding, 2016). Quantitative and qualitative components of mixed method studies were assessed separately (Kmet et al., 2004; Maharaj and Harding, 2016).

### Extracting the data

JA electronically extracted the names of authors, publication year, methodological approaches, participants, location, prescriber/referrers, referral pathways, facilities, intervention format, BSA type, facilitators, barriers, timescale/duration, dose/frequency, health conditions, and outcomes. Extracted data were inputted in a bespoke data collection form using Microsoft Excel.

### Synthesising the findings

We employed realist synthesis to analyse and synthesise extracted data (Pawson et al., 2005; Rycroft-Malone et al., 2012) about the facilitators, barriers, and descriptions of BPPs and how were they implemented. Using hybrid coding in NVivo 12 (Fereday and Muir-Cochrane, 2006), JA developed themes of contextual factors associated with BPP implementation. These contextual factors were used to develop the context-mechanism-outcome configurations elaborated by ‘if-then’ statements for the development of programme theories describing the mechanism of BPP implementation (Pawson et al., 2005; Rycroft-Malone et al., 2012; Pawson and Bellamy, 2006; Thomas and Harden, 2008). We developed programme theories for BPP implementation by refining the five initial social prescribing programme theories on patient enrolment, engagement, adherence, link worker coordination, and partnership with CBOs (Husk et al., 2020; Rycroft-Malone et al., 2012; Bickerdike et al., 2017; Chatterjee et al., 2018; Bertotti et al., 2018; Pawson and Bellamy, 2006). The developed themes of contextual factors and programme theories for BPP implementation were consulted and validated by JA in a series of virtual meetings and presentations with KH, KNI, SCh; members of the research advisory team (RH, SP, SCu); and other stakeholders from the Hydro Nation Forum, the policy governing body of the Hydro Nation Scholars Programme, which is the funding organisation of this research. The themes of contextual factors and programme theories were refined and finalised based on the comments from these consultations.

Our study was registered with PROSPERO (CRD42020170660).

## Results

### Search results

We collected 6,736 records from the database and snowball search (Figure 1). We excluded 3,814 duplicates before screening and 2,483 reports due to irrelevancy at title and abstract screening stage. We excluded 136 and 288 reports from database and citation searches, respectively, at full-text screening stage (Appendix 2).

**Figure 1.**
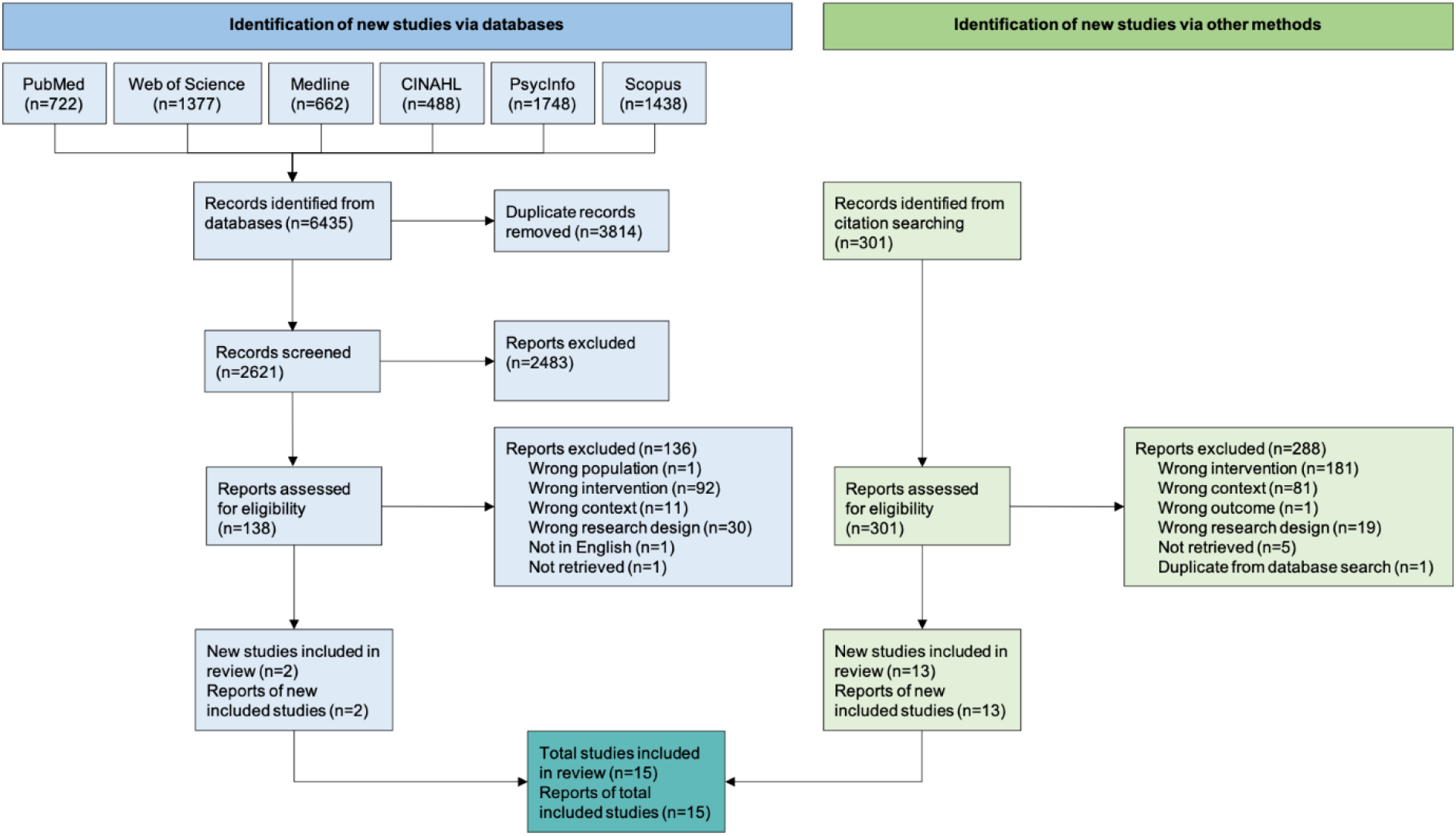
Study selection.

### Quality assessment results

We included 15 studies. Four quantitative studies had strong (Razani et al., 2018; Rogers et al., 2014; Vella et al., 2013; Maund et al., 2019), two had good (Godfrey et al., 2015; James et al., 2017), and three had adequate quality (de Matos et al., 2017; Hignett et al., 2017; Cavanaugh and Rademacher, 2014). Seven qualitative studies had adequate quality (Maund et al., 2019; White et al., 2016; Lopes, 2015; Bennett et al., 2014; Dustin et al., 2011; Fleischmann et al., 2011; Mowatt and Bennett, 2011). Quantitative papers were downgraded due to strategies on controlling confounders and reporting variance estimates. Qualitative papers were downgraded because of data collection, verification, and reflexivity procedures (Table 1) (Agarwal et al., 2013).

**Table 1.**
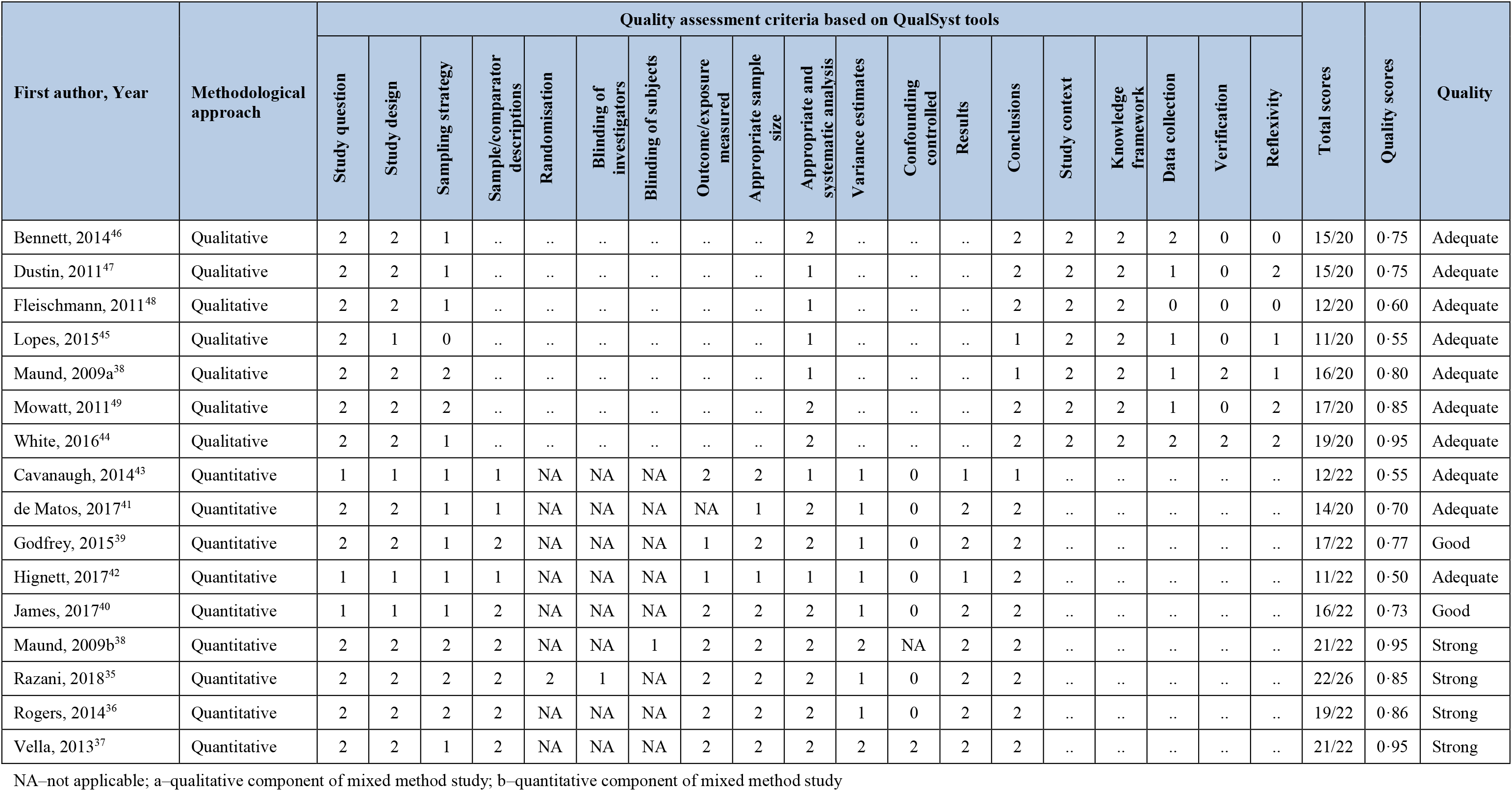
Results of quality assessment using QualSyst tools (reporting format adopted from Agarwal et al., 2013)

Since realist reviews require contextual configurations to describe mechanisms of programme implementation, we did not exclude studies which had below average quality results (Pawson et al., 2005; Rycroft-Malone et al., 2012).

### Characteristics of included studies

Eight studies were conducted in the USA (Razani et al., 2018; de Matos et al., 2017; Bennett et al., 2014; Rogers et al., 2014; Vella et al., 2013; Dustin et al., 2011; Fleischmann et al., 2011; Mowatt and Bennett, 2011), five studies in the UK (Maund et al., 2019; Godfrey et al., 2015; Hignett et al., 2017; Cavanaugh and Rademacher, 2014; White et al., 2016), and two in Portugal (de Matos et al., 2017; Lopes, 2015) (Table 2). Eight hundred three (803) service users aged between 2 and 85 years participated in the studies, of which 197 were veterans aged between 21 and 50 years. BSAs were referred to or prescribed by health, social care, and health-trained teachers. The prescription or referral was provided in healthcare (Razani et al., 2018; Rogers et al., 2014; Vella et al., 2013; Godfrey et al., 2015; James et al., 2017; White et al., 2016; Bennett et al., 2014; Dustin et al., 2011; Fleischmann et al., 2011; Mowatt and Bennett, 2011), social care (Maund et al., 2019; Godfrey et al., 2015; James et al., 2017; de Matos et al., 2017; White et al., 2016), and specialised educational facilities (Godfrey et al., 2015; Hignett et al., 2017; Cavanaugh and Rademacher, 2014) using SP referral pathways (Husk et al., 2020). In one study, all three facilities provided the referral or prescription (Godfrey et al., 2015).

**Table 2.**
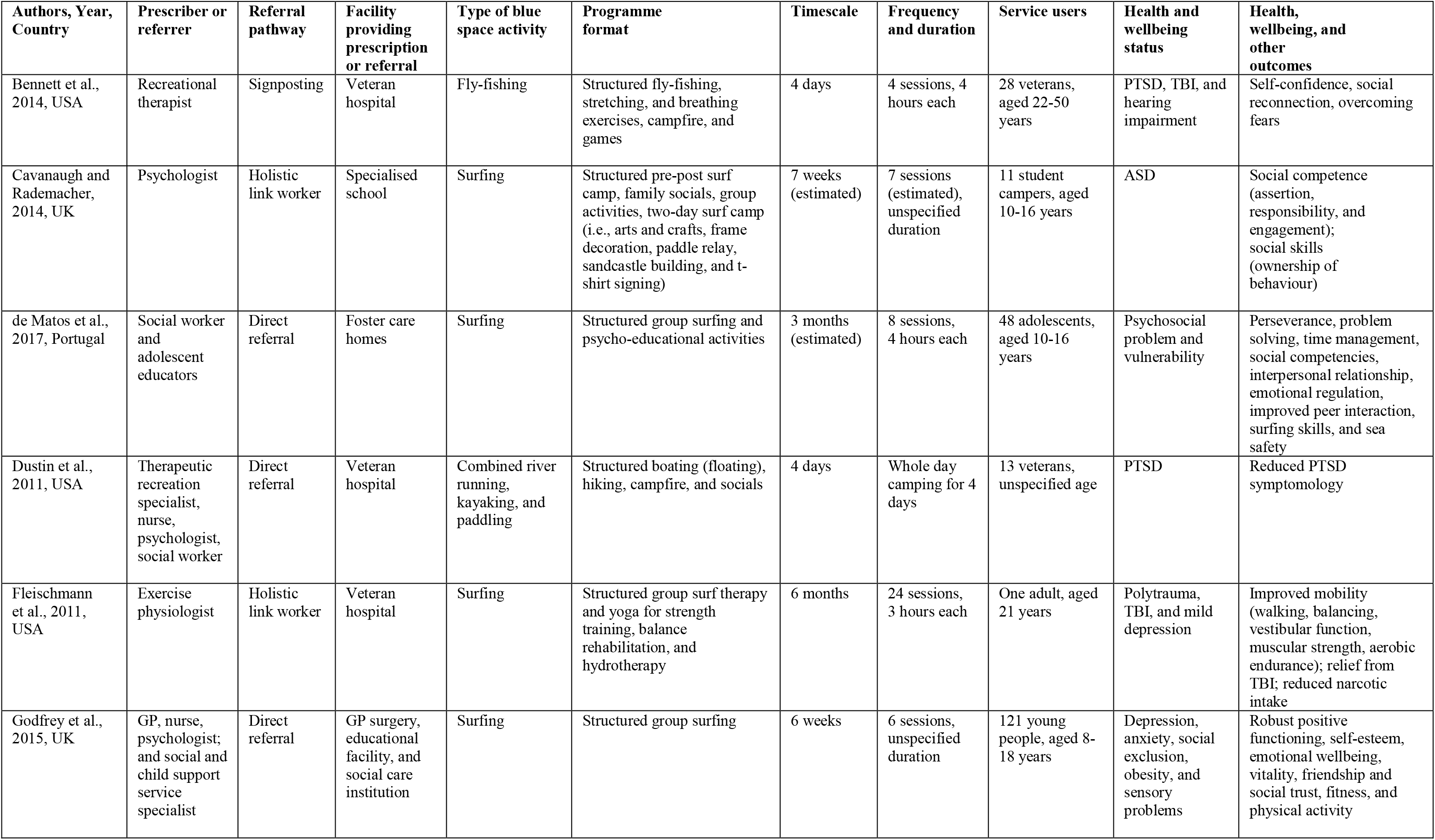
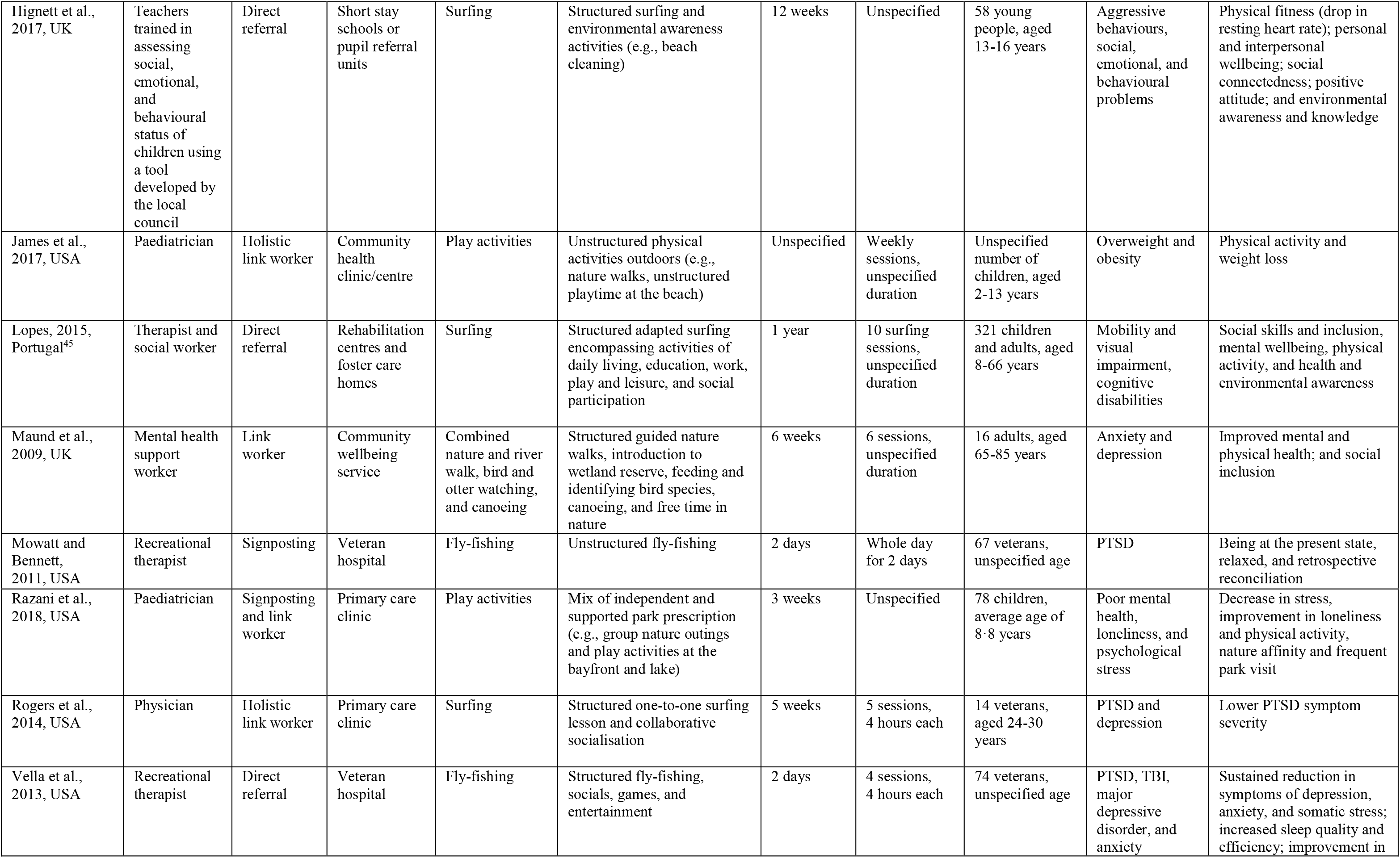
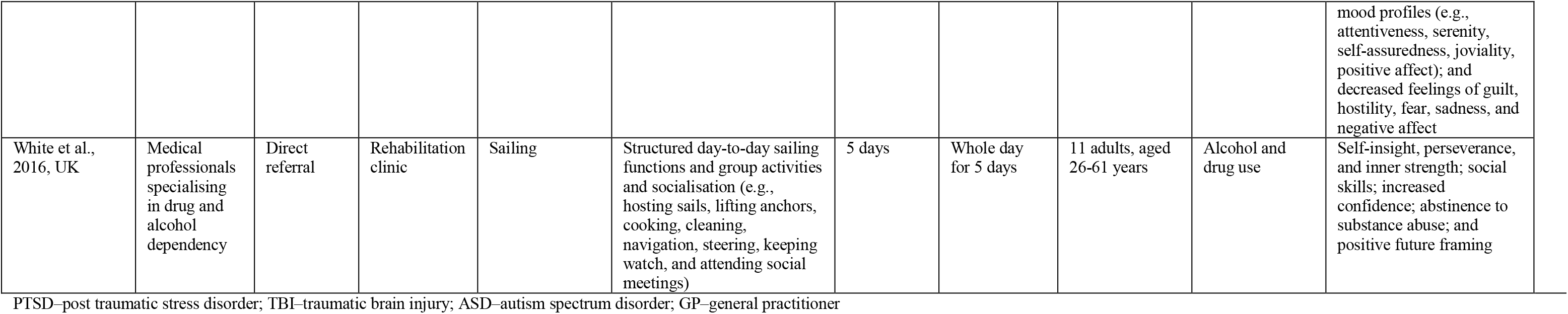
Characteristics of included studies and corresponding pre-existing social prescribing programme theories.

### Blue space activities prescribed in healthcare facilities

Ten studies were in military hospitals, GP practices, paediatric, and rehabilitation clinics (Razani et al., 2018; Rogers et al., 2014; Vella et al., 2013; Godfrey et al., 2015; James et al., 2017; White et al., 2016; Bennett et al., 2014; Dustin et al., 2011; Fleischmann et al., 2011; Mowatt and Bennett, 2011). Veterans with hearing impairment, anxiety, depression, TBI, and PTSD were prescribed with fly-fishing by recreational therapists through signposting or direct referral (Vella et al., 2013; Bennett et al., 2014; Mowatt and Bennett, 2011). Veterans with PTSD were directly referred to a combination of running, boating, kayaking, and paddling by a team of recreational therapist, nurse, psychologist, and social worker (Dustin et al., 2011). Some veterans with PTSD (Rogers et al., 2014) and children who were obese, had sensory problems, depression, anxiety, and PTSD (Godfrey et al., 2015) were prescribed with surfing by physicians (Rogers et al., 2014) or a team of GPs, nurses, and psychologists (Godfrey et al., 2015) through direct referral or holistic link worker. Socioeconomically deprived and ethnically diverse children with obesity, loneliness, and poor mental health were prescribed with play at a beach (Razani et al., 2018; James et al., 2017) by pediatricians using holistic link worker (James et al., 2017) or a combination of signposting and link worker pathways (Razani et al., 2018). Patients who experienced substance abuse were directly referred to sailing by substance abuse specialists (White et al., 2016). Participants had improvements in mood and emotions (Razani et al., 2018; White et al., 2016), better sleep quality (Vella et al., 2013), self-efficacy (White et al., 2016; Bennett et al., 2014; Mowatt and Bennett, 2011), reduced substance intake (White et al., 2016; Dustin et al., 2011), decreased stress and PTSD symptomatology (Rogers et al., 2014; Vella et al., 2013), and increased relaxation (Razani et al., 2018; Vella et al., 2013; Mowatt and Bennett, 2011). Some felt less depressed, anxious, and lonely (Razani et al., 2018) whilst others felt improvements in their mobility (Fleischmann et al., 2011), physical activity (Razani et al., 2018; James et al., 2017), and body weight (James et al., 2017). Others had improved social skills and increased contact with nature (Razani et al., 2018).

### Blue space activities prescribed in social care facilities

Four studies were in social care institution, foster care homes, and community wellbeing centre (Maund et al., 2019; Godfrey et al., 2015; de Matos et al., 2017; Lopes, 2015). Young people and adults with mobility and sensory impairment (Lopes, 2015), psychosocial problems, anxiety, and depression were directly referred to surfing by either a team of social worker and therapist (Lopes, 2015); social worker and adolescent educators (de Matos et al., 2017); or social child support specialist (Godfrey et al., 2015; de Matos et al., 2017; Lopes, 2015). Adults and elderly patients with anxiety and depression were referred to guided river walks, bird and otter watching, and canoeing by mental health workers through a link worker pathway (Maund et al., 2019). Participants improved daily functioning, self-esteem (Godfrey et al., 2015), fitness, physical activity (Godfrey et al., 2015; de Matos et al., 2017), mental and emotional wellbeing (Godfrey et al., 2015; de Matos et al., 2017), interpersonal competencies (Godfrey et al., 2015; de Matos et al., 2017; Lopes, 2015), and environmental knowledge (Lopes, 2015).

### Blue space activities prescribed in specialised educational institutions

Three studies were in specialised educational facilities and pupil referral units (Godfrey et al., 2015; Hignett et al., 2017; Cavanaugh and Rademacher, 2014). Young people with behavioural problems, ASD, depression, anxiety, and sensory problems were referred to surfing by psychologists, health-trained educators, or social and child support specialists using direct referral or holistic link worker pathways (Godfrey et al., 2015; Hignett et al., 2017; Cavanaugh and Rademacher, 2014). Participants had improvements in their attitude (Hignett et al., 2017), self-esteem (Godfrey et al., 2015), physical fitness (Godfrey et al., 2015; Hignett et al., 2017), mental and emotional wellbeing (Hignett et al., 2017), social competence (Godfrey et al., 2015; Hignett et al., 2017; Cavanaugh and Rademacher, 2014), and ecological knowledge (Hignett et al., 2017).

### Contextual factors and programmes theories describing the mechanisms of BPP implementation

The patterns of contextual factors (CFs) associated with the implementation of BPPs are either patient-related or programme-related. In total, we identified 20 patient and programme-related contextual factors (Table 3). The patient-related contextual factors are presentation of information on prescription or referral (CF1), patient perceptions of blue space activities (CF2), programme format (activity, timescale, frequency, duration) (CF3), accessibility (provision of transportation, equipment, other logistics) (CF4), compatibility of blue space activities to patient needs (CF5), social environment (CF6), blue space environment (CF7), skilled service providers (CF8), values of service providers (CF9), social support (CF10), and health, wellbeing, and other personal improvements of service users (CF11). The programme-related contextual factors are: coordination between link workers, service providers, and health and social care facilities (CF12), patient-to-link worker communication (CF13), patient-to-healthcare provider communication (CF14), patient-to-service provider communication (CF15), patient-to-patient communication (16), patient-to-carer communication (CF17), organisational support (staff and volunteers) (CF18), stakeholder support (CF19), and funding and policy support (CF20).

**Table 3.** Themes of contextual factors associated with BPP implementation and initial social prescribing programme theories.

| Source study | Identified contextual factors (CFs) | No. of initial social prescribing programme theories (PTs) supported by the study |
| --- | --- | --- |
| Bennett et al., 2014 | Programme format (activity, timescale, frequency, and duration) (CF3)<br>Accessibility (provision of transportation, equipment, and other logistics) (CF4)<br>Compatibility of blue space activities to patient needs (CF5)<br>Social environment (CF6)<br>Blue space environment (CF7)<br>Values of service providers (CF9)<br>Social support (CF10)<br>Health, wellbeing, and other personal improvements of service users (CF11)<br>Organisational support (staff and volunteers) (CF18)<br>Funding and policy support (CF20) | 1,2,3,5 |
| Cavanaugh and Rademacher, 2014 | Programme format (activity, timescale, frequency, and duration) (CF3)<br>Skilled service providers (CF9)<br>Health, wellbeing, and other personal improvements of service users (CF11)<br>Organisational support (staff and volunteers) (CF18) | 1,2,3,5 |
| de Matos et al., 2017 | Programme format (activity, timescale, frequency, and duration) (CF3)<br>Social environment (CF6)<br>Skilled service providers (CF9)<br>Social support (CF10)<br>Health, wellbeing, and other personal improvements of service users (CF11)<br>Coordination between link workers, service providers, and health and social care facilities (CF12)<br>Organisational support (staff and volunteers) (CF18) | 1,2,3,4,5 |
| Dustin et al., 2011, | Programme format (activity, timescale, frequency, and duration) (CF3)<br>Social environment (CF6)<br>Blue space environment (CF7)<br>Social support (CF10)<br>Health, wellbeing, and other personal improvements of service users (CF11) | 1,2,3,4,5 |
|  | Patient-to-patient communication (CF16)<br>Stakeholder support (CF19)<br>Funding and policy support (CF20) |  |
| Fleischmann et al., 2011 | Programme format (activity, timescale, frequency, and duration) (CF3)<br>Social environment (CF6)<br>Social support (CF10)<br>Health, wellbeing, and other personal improvements of service users (CF11)<br>Patient-to-patient communication (CF16) | 1,2,3,4 |
| Godfrey et al., 2015 | Programme format (activity, timescale, frequency, and duration) (CF3)<br>Accessibility (provision of transportation, equipment, and other logistics) (CF4)<br>Values of service providers (CF9)<br>Health, wellbeing, and other personal improvements of service users (CF11)<br>Organisational support (staff and volunteers) (CF18)<br>Funding and policy support (CF20) | 1,2,3,5 |
| Hignett et al., 2017 | Programme format (activity, timescale, frequency, and duration) (CF3)<br>Accessibility (provision of transportation, equipment, and other logistics) (CF4)<br>Blue space environment (CF7)<br>Health, wellbeing, and other personal improvements of service users (CF11)<br>Organisational support (staff and volunteers) (CF18)<br>Stakeholder support (CF19) | 1,2,3,5 |
| James et al., 2017 | Presentation of information on prescription or referral (CF1)<br>Programme format (activity, timescale, frequency, and duration) (CF3)<br>Accessibility (provision of transportation, equipment, and other logistics) (CF4)<br>Health, wellbeing, and other personal improvements of service users (CF11)<br>Coordination between link workers, service providers, and health and social care facilities (CF12)<br>Patient-to-link worker communication (CF13)<br>Patient-to-healthcare provider communication (CF14)<br>Patient-to-service provider communication (CF15)<br>Patient-to-carer communication (CF17)<br>Stakeholder support (CF19) | 1,2,3,4,5 |
| Lopes, 2015 | Patient perceptions of blue space activities (CF2)<br>Programme format (activity, timescale, frequency, and duration) (CF3)<br>Accessibility (provision of transportation, equipment, and other logistics) (CF4)<br>Compatibility of blue space activities to patient needs (CF5)<br>Social environment (CF6)<br>Blue space environment (CF7)<br>Values of service providers (CF9)<br>Social support (CF10) | 1,2,3,4,5 |
|  | Patient-to-patient communication (CF16)<br>Organisational support (staff and volunteers) (CF18)<br>Stakeholder support (CF19) |  |
| Maund et al., 2009 | Programme format (activity, timescale, frequency, and duration) (CF3)<br>Accessibility (provision of transportation, equipment, and other logistics) (CF4)<br>Social environment (CF6)<br>Blue space environment (CF7)<br>Values of service providers (CF9)<br>Social support (CF10)<br>Health, wellbeing, and other personal improvements of service users (CF11)<br>Organisational support (staff and volunteers) (CF18) | 1,2,3,5 |
| Mowatt and Bennett, 2011 | Social environment (CF6)<br>Blue space environment (CF7)<br>Values of service providers (CF9)<br>Social support (CF10)<br>Health, wellbeing, and other personal improvements of service users (CF11)<br>Patient-to-link worker communication (CF13)<br>Patient-to-healthcare provider communication (CF14)<br>Patient-to-service provider communication (CF15)<br>Patient-to-patient communication (CF16)<br>Patient-to-carer communication (CF17) | 2,3,4 |
| Razani et al., 2018 | Presentation of information on prescription or referral (CF1)<br>Programme format (activity, timescale, frequency, and duration) (CF3)<br>Accessibility (provision of transportation, equipment, and other logistics) (CF4)<br>Values of service providers (CF9)<br>Health, wellbeing, and other personal improvements of service users (CF11)<br>Patient-to-link worker communication (CF13)<br>Patient-to-healthcare provider communication (CF14)<br>Patient-to-service provider communication (CF15)<br>Patient-to-carer communication (CF17) | 1,2,3,4 |
| Rogers et al., 2014 | Patient perceptions of blue space activities (CF2)<br>Programme format (activity, timescale, frequency, and duration) (CF3)<br>Accessibility (provision of transportation, equipment, and other logistics) (CF4)<br>Social environment (CF6)<br>Values of service providers (CF9)<br>Social support (CF10)<br>Health, wellbeing, and other personal improvements of service users (CF11)<br>Patient-to-link worker communication (CF13) | 1,2,3,4 |
|  | Patient-to-healthcare provider communication (CF14)<br>Patient-to-service provider communication (CF15)<br>Patient-to-patient communication (CF16)<br>Patient-to-carer communication (CF17) |  |
| Vella et al., 2013 | Programme format (activity, timescale, frequency, and duration) (CF3)<br>Accessibility (provision of transportation, equipment, and other logistics) (CF4)<br>Social environment (CF6)<br>Blue space environment (CF7)<br>Social support (CF10)<br>Health, wellbeing, and other personal improvements of service users (CF11)<br>Organisational support (staff and volunteers) (CF18) | 1,2,3,5 |
| White et al., 2016 | Programme format (activity, timescale, frequency, and duration) (CF3)<br>Social environment (CF6)<br>Blue space environment (CF7)<br>Social support (CF10)<br>Values of service providers (CF9)<br>Health, wellbeing, and other personal improvements of service users (CF11) | 1,2,3 |

The combinations of 20 patient and programme-related contextual factors informed the development of programme theories describing the mechanisms of BPP implementation (Pawson et al., 2005; Rycroft-Malone et al., 2012; Pawson and Bellamy, 2006). Table 4 presents how the different contextual factors informed the development of final programme theories for BPP implementation based on the initial social prescribing programme theories.

**Table 4.** Final programme theories for BPP implementation using the identified contextual factors.

| Initial Social Prescribing Programme Theories (PTs) supported by included studies | Context | Setting | Contextual Factors (CF) | Final Programme Theories describing the mechanisms of implementing Bluespace Prescribing Programmes based on initial social prescribing programme theories |
| --- | --- | --- | --- | --- |
| Initial PT1: Patient enrolment<br>*If referral is presented in an acceptable manner, it is compatible with patient's need and expectations, and the patient believes that it will improve their condition, then they may enrol (Husk et al., 2020).* | People with physical and/or mental health conditions seek health and social care services | Health or social care facilities | CF1 Presentation of information on prescription or referral<br>CF2 Patient perceptions of blue space activities<br>CF3 Programme format (activities, timescale, frequency, and duration) | Final PT1: Patient enrolment to blue prescription programmes<br>*If service users' apprehensions and optimism are positively influenced by information on BPPs provided by prescribers, then they may enrol.* |
| Initial PT2: Patient engagement<br>*If transportation is provided making the socially prescribed activity accessible to the patient, then they will engage (Husk et al., 2020).* | People with physical and/or mental health conditions enrolled to a community-based blue prescription programme | Community-based organisation providing blue prescription programmes | CF3 Programme format (activity, timescale, frequency, and duration)<br>CF4 Accessibility (provision of transportation, equipment, and other logistics)<br>CF5 Compatibility of blue space activities to patient needs<br>CF6 Social environment<br>CF7 Blue space environment | Final PT2: Patient engagement with blue prescription activities<br>*If service users are provided with free logistics, equipment, and transportation to access a socially supportive and client-centred blue space activity and environment, then they may engage.* |
| Initial PT3: Patient adherence<br>*If the service providers are skilled and there are improvements on patient's condition, then they are more likely to keep attending (Husk et al., 2020).* | People with physical and/or mental health condition engaging with community-based blue space activities | Community-based organisation providing blue prescription programmes | CF8 Skilled service providers<br>CF9 Values of service providers<br>CF10 Social support<br>CF11 Health, wellbeing, and other personal improvements of service users | Final PT3: Patient adherence to blue prescription programmes<br>*If service users experience social support and health improvement through blue space activities delivered by knowledgeable and skilled service providers, then they may adhere.* |
| Initial PT4: Link worker coordination<br>*If there are link workers facilitating delivery of social prescriptions and liaising with health and social care facilities and third sector organisations, then the referral uptake will be successful (Husk et al., 2020; Bickerdike et al., 2017; Chatterjee et al., 2018; Bertotti et al., 2018).* | People with physical and/or mental health conditions seek health and social care (some were accompanied by their carers) | Health or social care facilities and community-based organisations | CF12 Coordination between link workers, service providers, and health and social care facilities<br>CF13 Patient-to-link worker communication<br>CF14 Patient-to-healthcare provider communication<br>CF15 Patient-to-service provider communication<br>CF16 Patient-to-patient communication<br>CF17 Patient-to-carer communication | Final PT4: Visibility of blue prescription programmes in health and social care facilities and successful prescription supported by communication protocols<br>*If communication protocols are in place between service users, prescribers, link workers, and service providers, then BPPs may be more visible and successfully implemented in health and social care facilities.* |
| Initial PT5: Partnership with community-based organisations<br>*If partnership between health and social care facilities and community-based organisations is financially supported, then the delivery of social prescribing programmes will be sustained (Bickerdike et al., 2017; Chatterjee et al., 2018; Bertotti et al., 2018).* | People with physical and/or mental health conditions seek health and social care (some are accompanied by their carers) | Health or social care facilities and community-based organisations | CF18 Organisational support (staff and volunteers)<br>CF19 Stakeholder support<br>CF20 Funding and policy support | Final PT5: Long-term programme sustainability of blue prescription programmes<br>*If BPPs are supported by organisational, stakeholder, funding, and policy interventions, then these are more likely to be sustainably implemented.* |

Figure 2 illustrates the delivery of BPP in health and social care settings and at what stage or component of the implementation where each programme theory is associated.

**Figure 2.**
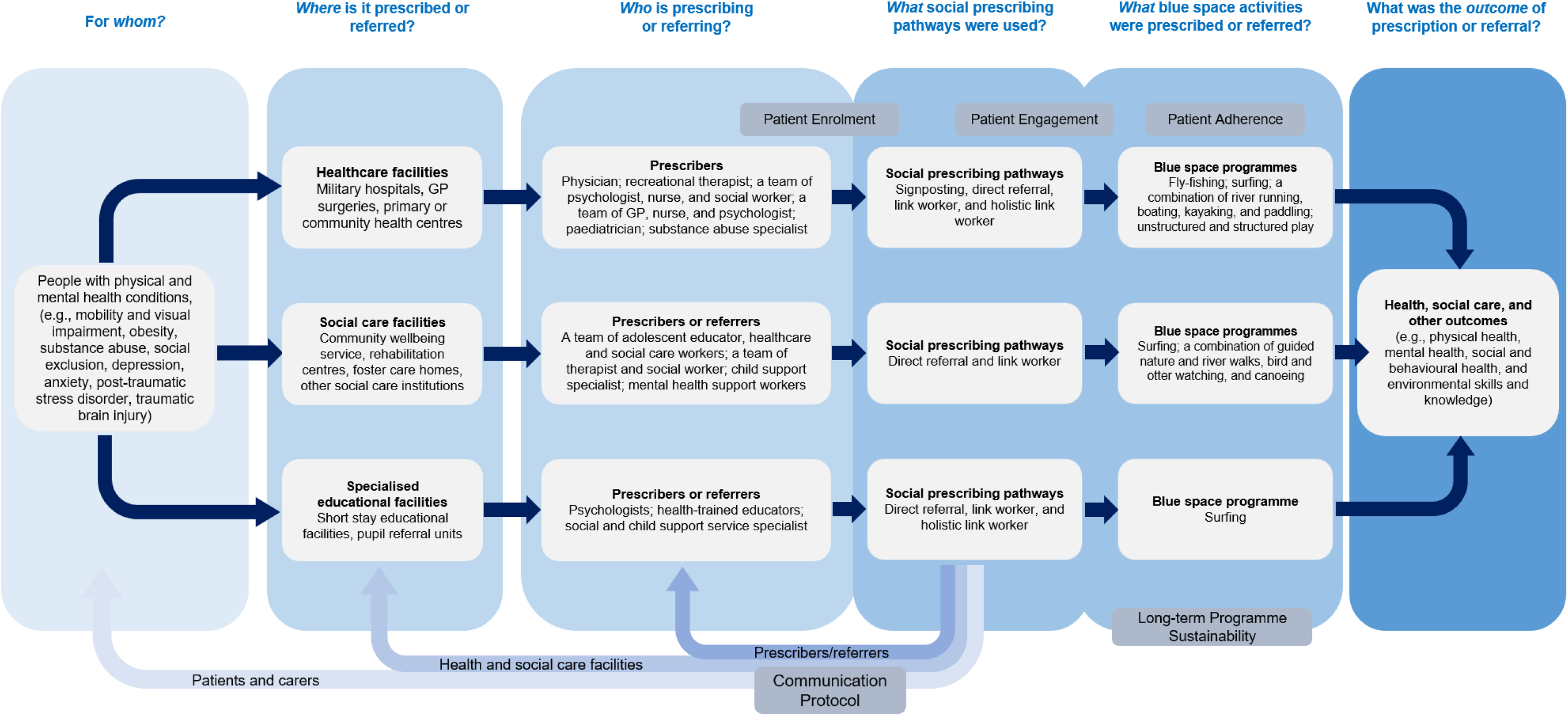
Context and mechanism of prescribing blue space activities in health, social care, and specialised educational facilities.

### Final Programme Theory 1: If service users’ apprehensions and optimism are positively influenced by information on BPPs provided by prescribers, then they may enrol

**Figure 3.**
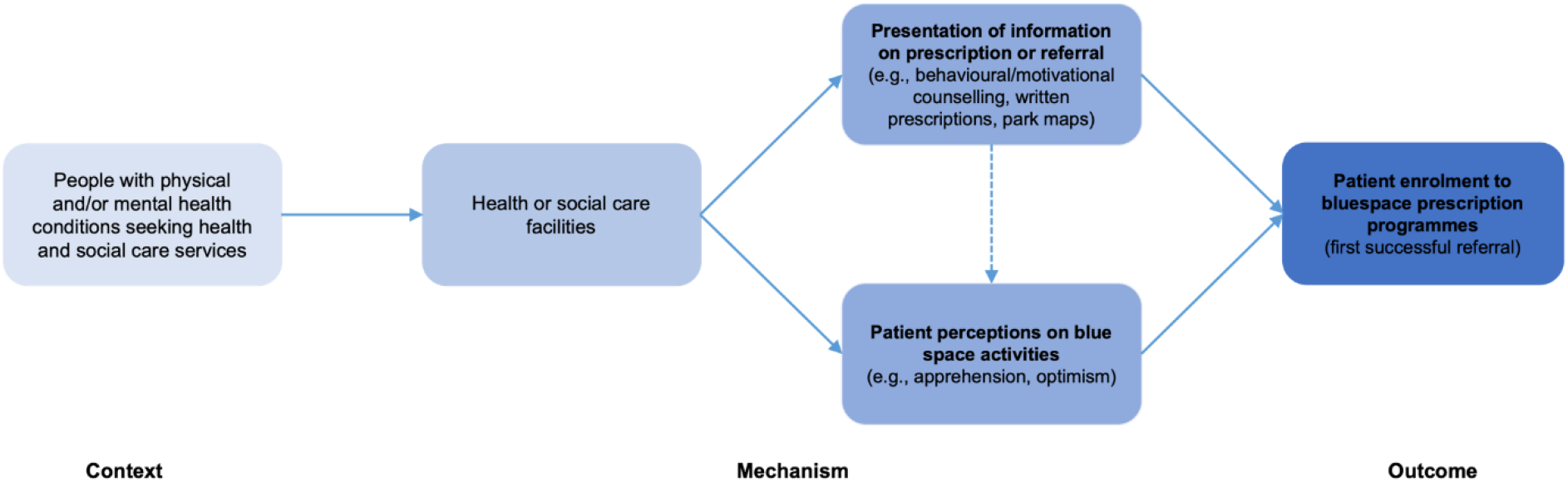
Contextual configurations for patient enrolment programme theory.

Some service users were optimistic towards BPPs, whilst others had apprehensions (Rogers et al., 2014; Lopes, 2015). The novelty of BPPs, unfamiliar environments, and patient’s lack of experience generated fear and anxiety (Lopes, 2015). Knowledge sharing through counselling using information about the type, structure, and benefits of BPPs, coupled with maps, pedometers, and activity guides informed patients’ decisions to enrol (Razani et al., 2018; James et al., 2017). Patients’ enrolment was guided by written prescriptions and initiated contact by link workers (James et al., 2017). Some healthcare workers raised concerns about filling out paper prescriptions being time-consuming and suggested integrating prescriptions of BSAs in the electronic prescribing system (James et al., 2017). Structured BSAs followed programmes of activities (Rogers et al., 2014; Vella et al., 2013; Maund et al., 2019; Godfrey et al., 2015; James et al., 2017; de Matos et al., 2017; Hignett et al., 2017; Bennett et al., 2014) whilst others were provided in an unstructured manner (Razani et al., 2018; James et al., 2017; Mowatt and Bennett, 2011). BPPs were provided as stay-in which required free meals and lodging (Vella et al., 2013; Hignett et al., 2017; Bennett et al., 2014; Dustin et al., 2011; Mowatt and Bennett, 2011), or stay-out (Razani et al., 2018; Rogers et al., 2014; Maund et al., 2019; Godfrey et al., 2015; James et al., 2017; White et al., 2016; Lopes, 2015; Fleischmann et al., 2011).

### Final Programme Theory 2: If service users are provided with free logistics, equipment, and transportation to access a socially supportive and client-centred blue space activity and environment, then they may engage

**Figure 4.**
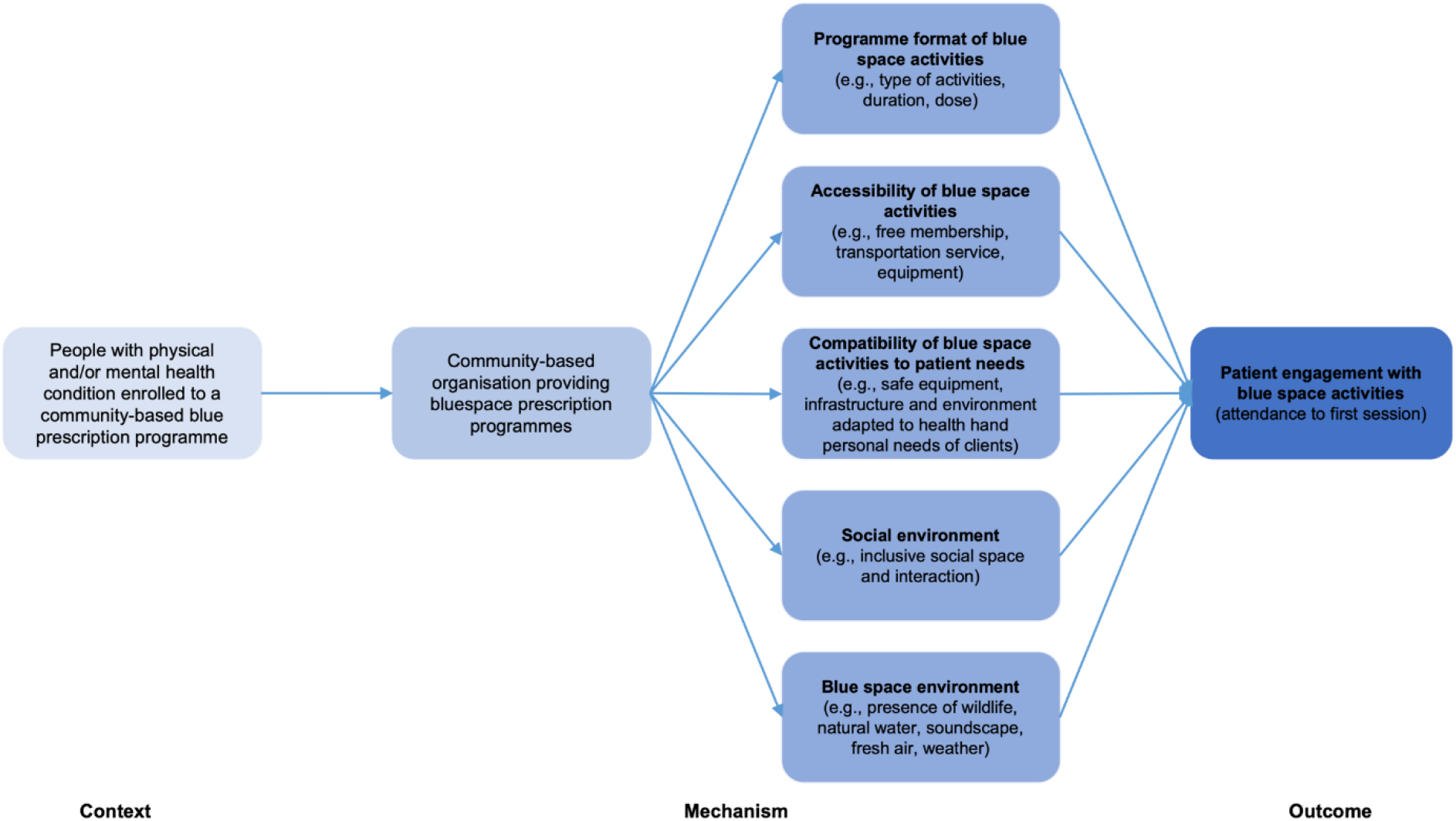
Contextual configurations for patient engagement programme theory.

Patient engagement was associated with patient preference, skills, and psychosocial fulfilment after initial sessions (Godfrey et al., 2015; Hignett et al., 2017). Consulting people with disabilities about the design of safety and supportive infrastructure making the environment accessible and adaptive to their needs enabled engagement (Lopes, 2015; Bennett et al., 2014). Service users had strong interests in BPPs because these were free (James et al., 2017). However, some BPPs (surfing, canoeing, kayaking) required costly equipment, especially for economically deprived service users. Providing equipment, transportation, and camping fees encouraged engagement especially for activities in distant locations (Razani et al., 2018; Rogers et al., 2014; Vella et al., 2013; Maund et al., 2019; Godfrey et al., 2015; James et al., 2017; Bennett et al., 2014). Travelling in groups to new places also relieved anxious participants (Maund et al., 2019). Wildlife and natural blue space or water environments facilitated relaxing experiences, distraction, engagement of human senses, self-strength, and acclimatisation (Vella et al., 2013; Maund et al., 2019; White et al., 2016; Bennett et al., 2014; Dustin et al., 2011). Weather was an environmental effect modifier (White et al., 2020; Hignett et al., 2017).

### Final Programme Theory 3: If service users experience social support and health improvement through blue space activities delivered by knowledgeable and skilled service providers, then they may adhere

**Figure 5.**
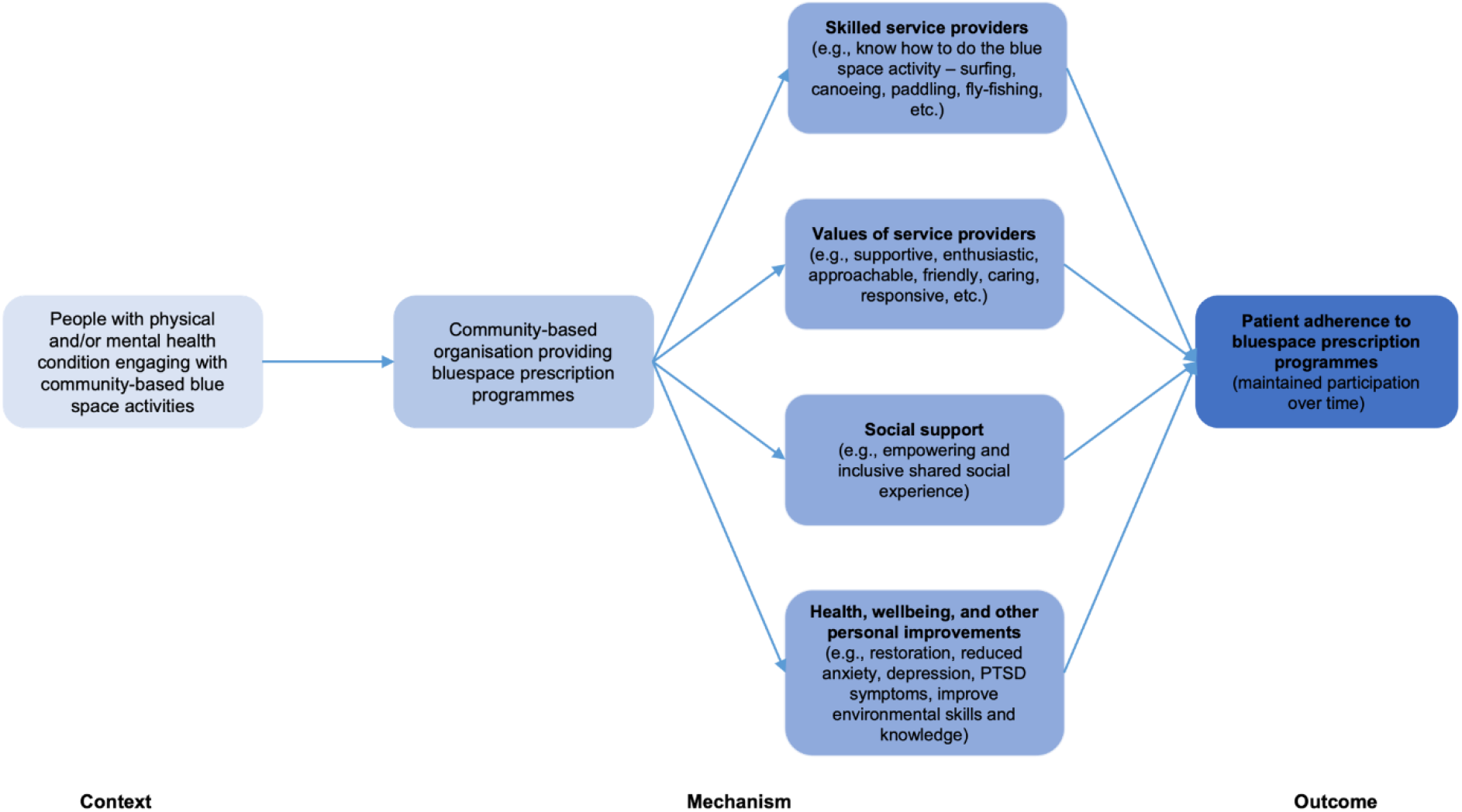
Contextual configurations for patient adherence programme theory.

Skilled service providers ensured standardised programme delivery using regularly evaluated guidelines (Hignett et al., 2017). Service providers were also trained on working with clients with disabilities (proper use of surfing boards for children with ASD) (Cavanaugh and Rademacher, 2014; Lopes, 2015). Positive values of service providers were appreciated by participants (Razani et al., 2018; Rogers et al., 2014; Maund et al., 2019; Hignett et al., 2017; White et al., 2016; Lopes, 2015; Bennett et al., 2014). Encouragement, enthusiasm, and positive reinforcement through constructive observations created a friendly atmosphere and empowered participants to explore new activities (Razani et al., 2018; Rogers et al., 2014; Maund et al., 2019; White et al., 2016; Lopes, 2015). Staff responsiveness facilitated connection with participants (Mowatt and Bennett, 2011) through open communications (Razani et al., 2018). This was perceived by clients as genuine care and willingness to help (Rogers et al., 2014). Improvements in environmental awareness and knowledge (Hignett et al., 2017; Lopes, 2015), self-esteem (Rogers et al., 2014; Vella et al., 2013; Maund et al., 2019; Godfrey et al., 2015; Cavanaugh and Rademacher, 2014; White et al., 2016; Lopes, 2015), mental health (Razani et al., 2018; Rogers et al., 2014; Vella et al., 2013; Maund et al., 2019), physical health (Maund et al., 2019; Hignett et al., 2017; Fleischmann et al., 2011), social health (de Matos et al., 2017; Hignett et al., 2017; White et al., 2016), and reduced dependency on medicines (Dustin et al., 2011; Fleischmann et al., 2011) and addictive substances (White et al., 2016) were also associated with patient adherence.

### Final Programme Theory 4: If communication protocols are in place between service users, prescribers, link workers, and service providers, then BPPs may be more visible and successfully implemented in health and social care facilities

**Figure 6.**
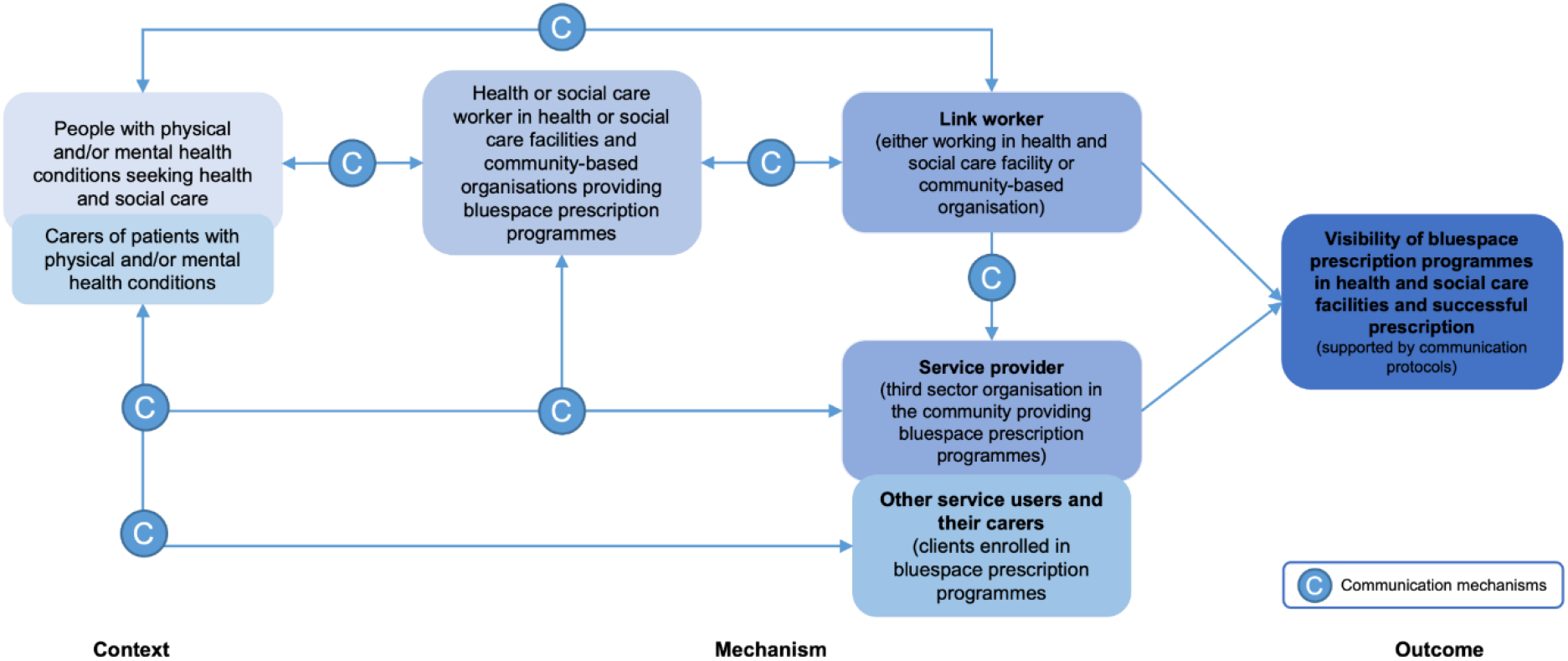
Contextual configuration for communication protocol programme theory.

Different communication channels were used within and outside health and social care facilities. Initial presentation of BPPs to prescribers facilitated planning of referral processes, requirements, and responsibilities (James et al., 2017; de Matos et al., 2017). Electronic communications provided feedback channels to discuss patients’ health conditions (James et al., 2017). Champions or advocates of BPPs in health and social care settings acted as programme leads and maintained coordination and programme visibility in health and social care facilities (James et al., 2017). Inclusive communication between health and social care workers, patients, and service providers facilitated by link workers and considered patient-preferred mechanisms and related attributes (Razani et al., 2018; James et al., 2017; Mowatt and Bennett, 2011) were also associated with health improvements (Rogers et al., 2014). Communications between patients and carers provided spaces for awareness raising and socialisation (Lopes, 2015). Socialisation between patients through interest-based grouping and matching also contributed to self-improvement (Razani et al., 2018; Rogers et al., 2014; James et al., 2017; de Matos et al., 2017; Lopes, 2015; Mowatt and Bennett, 2011).

### Final Programme Theory 5: If BPPs are supported by organisational, stakeholder, funding, and policy interventions, then these are more likely to be sustainably implemented

**Figure 7.**
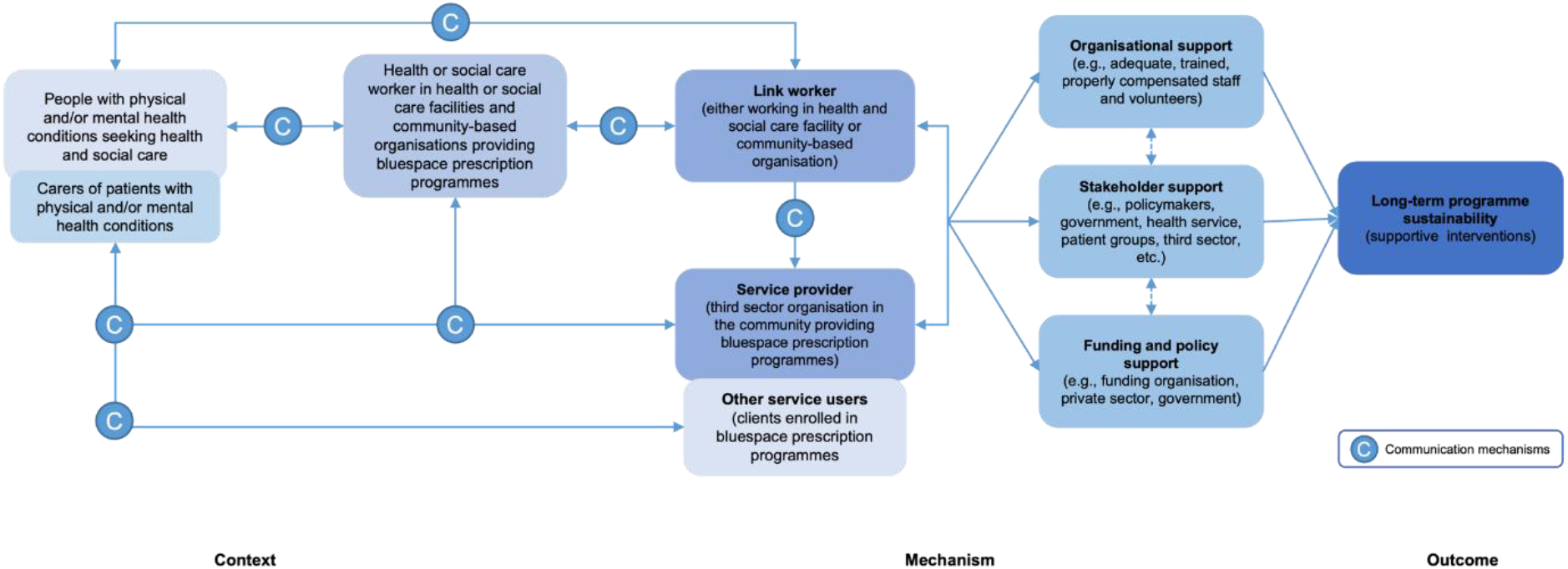
Contextual configuration for long-term programme sustainability programme theory.

Surfing was perceived as a beneficial health and social care intervention by health sector stakeholders, general public, and policymakers (Lopes, 2015). The health and social benefits of BPPs convinced funders, councils, and schools to support the programme (Hignett et al., 2017), especially because the cost of therapeutic surfing (£50/session) was less than the mental healthcare cost for young people (£265/year) (de Matos et al., 2017; Knapp et al., 2011). However, this economic evaluation was not robust (de Matos et al., 2017). Many BPPs were created through health and CBO partnerships (James et al., 2017; Dustin et al., 2011). Other BPPs received financial support from funding institutions (Godfrey et al., 2015; Hignett et al., 2017). Shared resources covered many implementation aspects (e.g., meals, transportation, equipment) making BPPs accessible to service users (Godfrey et al., 2015; James et al., 2017; Hignett et al., 2017; Lopes, 2015; Bennett et al., 2014). Adequate skilled staff and volunteers were the backbones of BPPs. Client-provider ratio depended on participant intake, activities, and financial capacity (Maund et al., 2019; Godfrey et al., 2015; James et al., 2017; de Matos et al., 2017; Cavanaugh and Rademacher, 2014).

## Discussion

Bluespace Prescription Programme (BPP) implemented in health and social care facilities contextualised by programme theories on patient enrolment, engagement, adherence, communication protocol, and programme sustainability had benefits on general, physical, mental, social health, and ecological knowledge of service users. These programme theories were shaped by configurations of contextual factors on patient needs and characteristics, accessibility, compatibility, social and blue space environments, skills and values of service providers, patient health improvement, communication, multi-stakeholder partnership, financing, and policy.

Outcomes investigated in this review are consistent with current evidence demonstrating that exposure to blue spaces has physical and mental health benefits (Völker and Kistemann, 2011; Gascon et al., 2017). Additionally, some BPPs had educational activities about the environment and these improved participants’ knowledge about the value of blue spaces and paved way for the development of pro-environmental behaviours (White et al., 2020). BPPs package the health-promoting impacts of blue spaces into structured health and social care programmes for those who could most benefit from these. From a behaviour change perspective (Michie et al., 2011), improving the quality and accessibility of blue spaces could provide greater opportunities for contact with these environments (White et al., 2020). However, these environmental modifications should be coupled with capability-building (knowledge sharing) and motivational strategies (social support) to promote and sustain contact with natural settings (Sharma et al., 2020), such as blue spaces.

BPPs target individuals’ motivations to increase utilisation of blue spaces for physical or social activities (Depledge and Bird, 2009; Britton et al., 2020). However, BPPs might not be for everyone as environmental and personal circumstances are associated with engagement (White et al., 2020). Medicalisation of spending time in nature could be perceived as a medical order rather than a personal choice (Tester-Jones et al., 2020). Prescribing specific blue spaces could reduce patients’ intrinsic motivation to visit nature, thus compromising its health benefits (Tester-Jones et al., 2020). Patient autonomy (Varkey, 2021) through a patient-centred approach using motivational interview should be considered in selecting BSAs which are compatible with patients’ health and personal needs (Britten et al., 2000; Little et al., 2001; Cockburn and Pit, 1997). The investigated BPPs in this review had higher duration than the 120 minutes per week recommended dose of nature (White et al., 2019). However, duration could be a modifying factor for nature engagement (White et al., 2020). Duration of BPPs should be matched with patients’ gender (Alejandre and Lynch, 2020; Vert et al., 2019; Elliott et al., 2018), age (Alejandre and Lynch, 2020; Garrett et al., 2019; Helbich et al., 2019; Dempsey et al., 2018), ethnicity (Leeworthy, 2001; Wolch and Zhang, 2004), and health conditions (Wood et al., 2016; Amoly et al., 2014). Increasing choices for BSAs such as dragon boating (Britton et al., 2020), recreational SCUBA diving (Dimmock, 2009), open water swimming (van Tulleken et al., 2018; Huttunen et al., 2004), and cycling near blue spaces (Roberts et al., 2018; Mackay and Neill, 2010; Pretty et al., 2007) could promote inclusivity and attract enrolment and engagement of many individuals with different personal characteristics and health needs. Engagement with and adherence to BPPs could be facilitated by positive doctor-patient interaction because it promotes patient’s self-confidence, motivation, and optimism (Ha and Longnecker, 2010). However, in one systematic review, only six studies had healthcare professionals prescribing BSAs (Britton et al., 2020). Some BSAs are prescribed by a team of health and social care workers with different backgrounds, specialisations, and expeertise. Appropriateness training of prescribers is an SP barrier (Britton et al., 2020), thus, health and social care professionals with varied backgrounds and skill sets should be provided with needs-based and standardised trainings on delivery protocols.

Realist reviews on general social prescribing and greenspace programmes highlighted doctor-patient relationship, capacity of service providers, and supportive social environments as contextual factors to implementation (Husk et al., 2020; Masterton et al., 2020), these were also highlighted in our review. Additionally, communication protocols, stakeholder collaboration, and policy support are fundamental programme-related factors associated with programme continuity (Masterton et al., 2020; McHale et al., 2020). Communication protocols between prescribers, link workers, and service providers facilitated by written prescriptions were essential elements of the referral pathways (Husk et al., 2020). England’s National Health Service backs the link worker-based model (NHS England, 2019) because link workers support healthcare delivery by facilitating resolutions for patient’s health concerns (Husk et al., 2020; Bertotti et al., 2018) through motivational counselling and by coordinating referrals between prescribers, patients, and service providers (Bickerdike et al., 2017; Chatterjee et al., 2018; Bertotti et al., 2018). Moreover, written prescriptions increased likelihood of patient enrolment because these were perceived as alternative to pharmaceutical prescriptions (McHale et al., 2020; Dempster et al., 2015). Many BPPs are also formed through health and stakeholder partnerships which are funding dependent. Unstable funding affects sustained SP organisation and delivery (Bertotti et al., 2018). There is also a perceived risk in data sharing between healthcare facilities and CBOs. Policies on institutionalising BPPs in government-funded health services and data sharing agreement between stakeholders are necessary. Moreover, medicalising ecosystem services of blue spaces could go against some ecological paradigms (Ofiara and Brown, 1999) within the scope of the environment sector. BSAs could cause disturbances to fish and other wildlife, human-induced pollution, and other irreversible ecosystem degradation (Maxwell et al., 2018; Burgin and Hardiman, 2011; Graham and Cooke, 2008; Davenport and Davenport, 2006). The ethical principles of beneficence and nonmaleficence (Varkey, 2021) should be extended for the protection of nature by involving healthcare specialists, environmentalists, patient groups, and blue space communities in planning and implementing BPPs that aims for the protection of patients and the environment.

In the context of the COVID-19 pandemic, the health-related use of outdoor environments increased during lockdowns (Hartig et al., 2014; Hubbard et al., 2021; Geng et al., 2021) suggesting that outdoor settings such as blue spaces helped people cope with stress (Heo et al., 2020). People in Scotland and Spain associated exposure to nature with mental health improvements and better sleep quality (Hartig et al., 2014; Corley et al., 2021). Contact with nature is associated with fewer mental and physical symptoms and may buffer the impacts of social isolation (Yang et al., 2021; Bratman et al., 2019; Van den Berg et al., 2015). BPPs could be used as an additional healthcare service to help the health sector meet increasing demands for mental healthcare. Link workers could facilitate BPPs through virtual health assessments and motivational interviewing (Razai et al., 2020). However, accessibility, safety, and financial concerns were also associated with less time and fewer visits in blue spaces (Astell-Burt and Feng, 2021) which should be considered and resolved. A siloed approach in tackling the mental health epidemic compounded by lockdowns and social isolation will not work as we transition to a ‘better normal’ after the COVID-19 pandemic. Approaches should include strong collaboration between health, environment, urban planning, third sector, amongst others to share more resources toward integrating BPPs in primary healthcare with appropriate environmental-economic accounting (United Nations-Food and Agriculture Organisation, European Union, Organisation for Economic Co-operation and Development, and World Bank Group, 2014). These collaborations and resources should ensure that contextual factors influencing enrolment, engagement, and adherence to BPPs are addressed along with effective communication and programme continuity.

### Strengths and limitations

We did not include grey literature and we only focused on BPPs prescribed in health and social care settings. Reports on BPPs in other facilities which are unpublished or not peer-reviewed could provide more information about other contextual factors and mechanisms of implementation. There were also limitations on the quality of individual studies. Many quantitative studies had quality issues on the strategies for controlling confounders and reporting variance estimates, whilst qualitative papers had issues on strategies for data collection, verification, and reflexivity.

Certain limitations are also anchored in employing a realist review. Realist synthesis is interpretative and subjective in nature (Masterton et al., 2020), especially in interpreting how contextual factors inform the development of programme theories. We ensured to remove this subjectivity bias by validating the results of the analysis during a series of stakeholders’ meetings and consultation with a multidisciplinary set of co-authors that informed the finalisation of the programme theories for BPP implementation. We were also limited on the available information about the contexts and mechanisms of each bluespace prescription programme in each study, especially when there is no detailed information about the intervention.

Geographical homogeneity was also a limitation. Some studies had participants from different ethnicities and sociodemographic backgrounds, but all studies were conducted in the Global North. Nonetheless, our study offers pragmatic recommendations for policy development supporting BPP implementation in different contexts. In the context of Global South countries, it is important to be cognisant of the political and social constructs surrounding blue space accessibility and the readiness of health systems for large-scale BPP implementation. Nevertheless, universal healthcare reforms in developing countries could leverage the groundwork on BPP integration in health promotion strategies to deliver the vision of healthy, liveable, and sustainable environments for all.

### Research directions

We suggest the conduct of intervention studies examining the intersection of BPPs, physical, and mental health as cost-effective programmes. Implementation research and pilot studies are needed to establish the ‘proof of concept’ of BPPs as a viable public health intervention for scaling-up and institutionalisation in national and local health and social care systems. Causation and long-term implications of BPPs on population health, healthcare delivery, and the environment should be investigated through randomised controlled trials and longitudinal studies.

## Conclusion

In summary, this realist review demonstrates that health, social care, and health-trained professionals working in health, social care, and specialised educational facilities provided referral to or prescription of BSAs. Service users of BPPs were veterans, adults, older people, and children who experienced physical and mental health conditions, social isolation, addiction, and/or behavioural issues. BPPs improved the physical, mental, and social health, as well as environmental knowledge of service users. The implementation of BPPs is associated with 20 unique patient and programme-related contextual factors on patient needs and characteristics, accessibility, compatibility, social and blue space environments, skills and values of service providers, patient health improvement, communication, multi-stakeholder partnership, financing, and policy. The configurations of these contextual factors informed the refinement of pre-existing social prescribing programme theories for the development of programme theories of BPPs. If implementation of accessible and patient centred BPPs is sustainably supported by multi-stakeholder partnerships, funding, policy interventions, effective communication protocols, skilled health and social care workers and service providers, then service users are more likely to enrol in, engage with, adhere to, and have improvements to health and environmental knowledge after the programme. Contextual factors and programme theories of investigated BPPs could inform development and implementation of similar nature-based social prescribing programmes in other contexts. BPPs could be an additional health and social care service that promote exposure to blue spaces for public health promotion and health improvement of individuals with multiple and long-term health conditions. With inaccessibility, long waiting list, adverse effects, incompatibility, cost, and environmental impacts of conventional healthcare, integrating patient centred and sustainable BPPs into a suite of healthcare services is beneficial.

## Data Availability

Beginning three months after publication until September 2023, database records and extracted data from included studies will be electronically shared to researchers who provide a methodologically sound proposal sent to the corresponding author. The request will be approved by the corresponding author and the director of studies.

## Acknowledgments

The corresponding author acknowledges funding from the Hydro Nation Scholars Programme of the Scottish Government. Author KNI [retracted for anonymity] acknowledges funding from the Scottish Government’s Rural and Environment Science and Analytical Services Division (RESAS). We would like to thank the Blue-Green Prescribing Reviewers Group composed of 11 researchers: A Estandarte (France); A San Buenaventura, MLA Vega, MAR Manalili, PJ Algones (Philippines); A Konapur, R Ungarala, S Medithi (India); C Lewnet, K Munro (United Kingdom); and V Ashipala (Namibia).

---

**Appendix 1.**
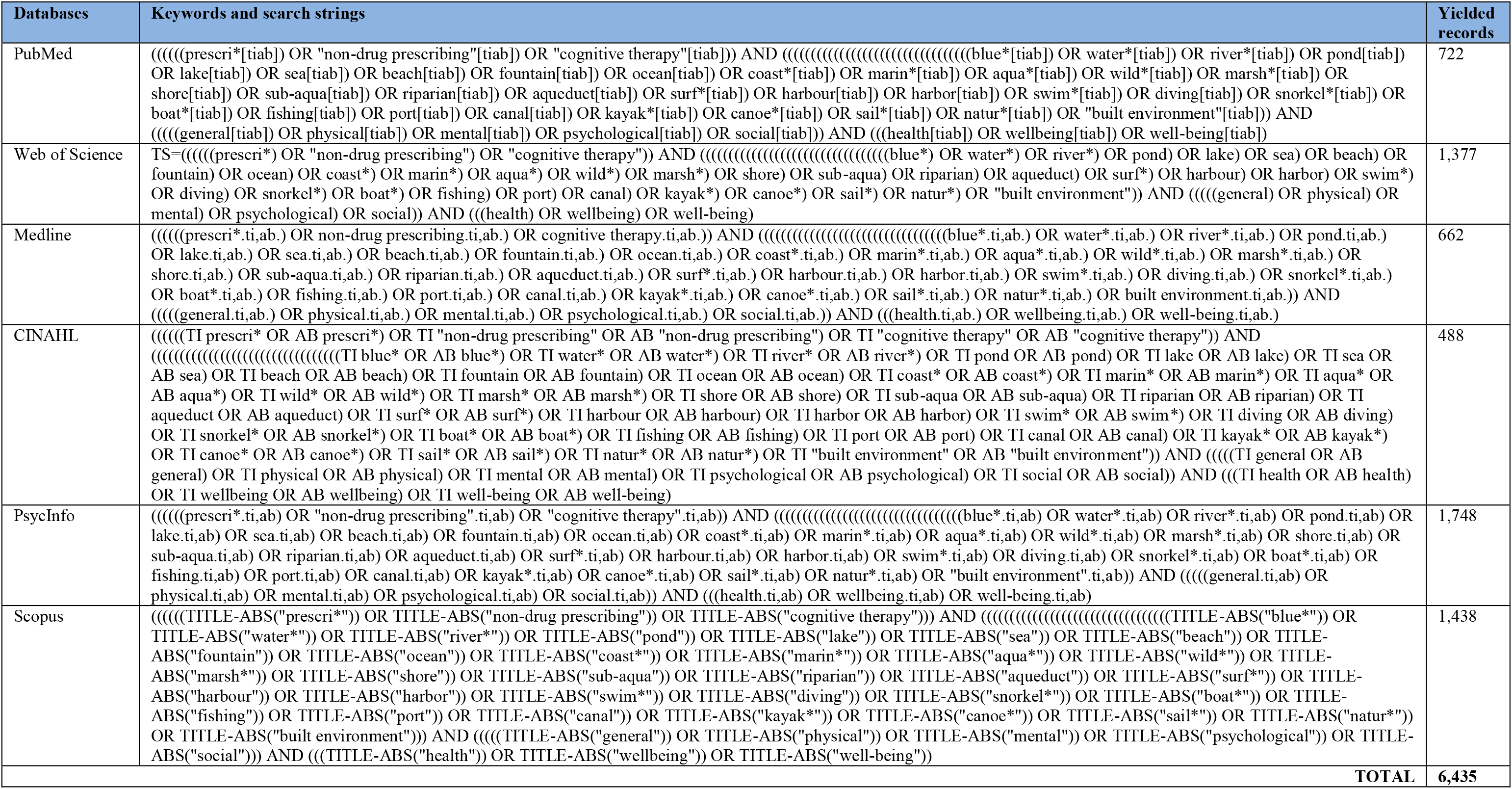
Keywords, search strings, and yielded records from database search.

**Appendix 2.**
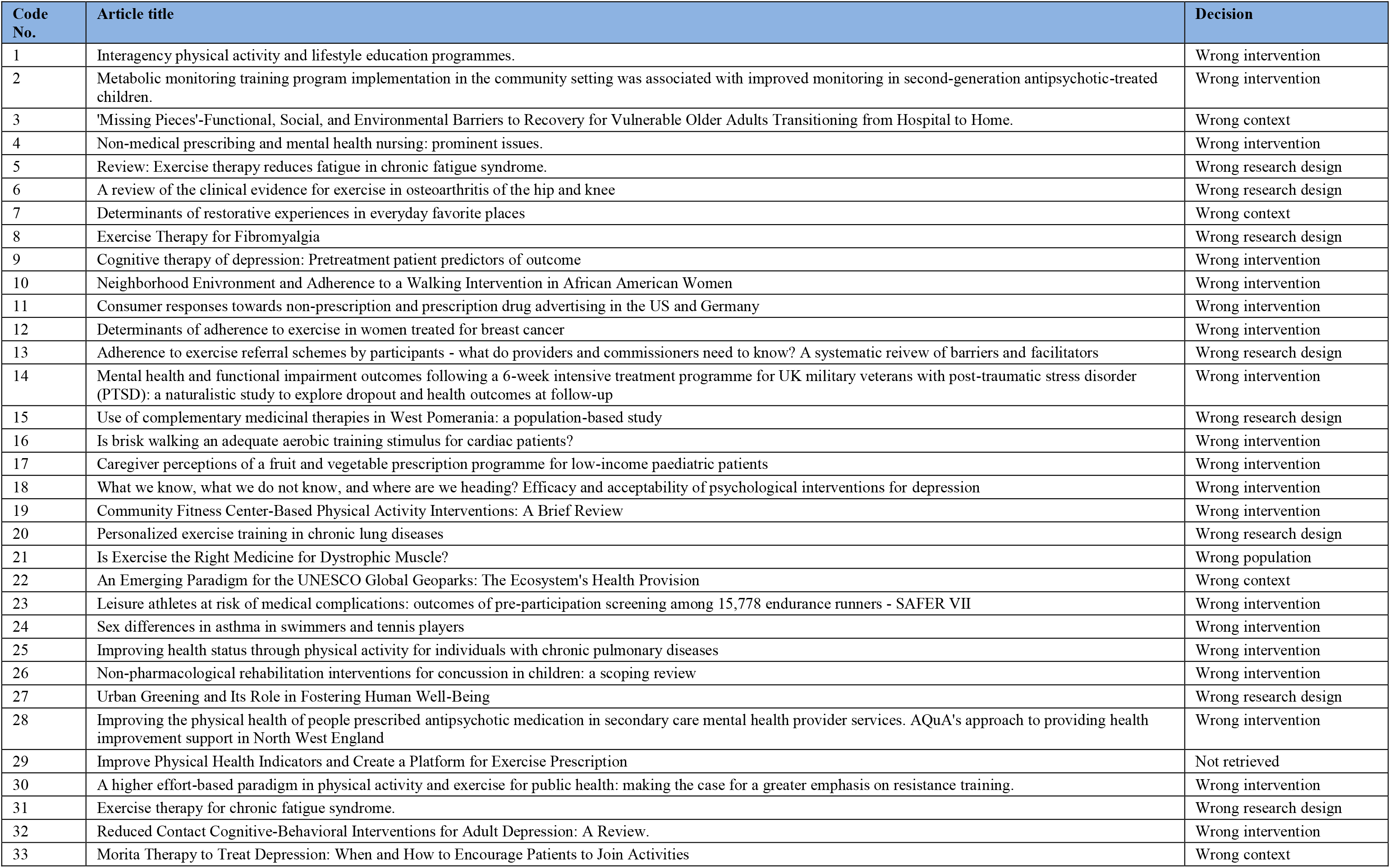
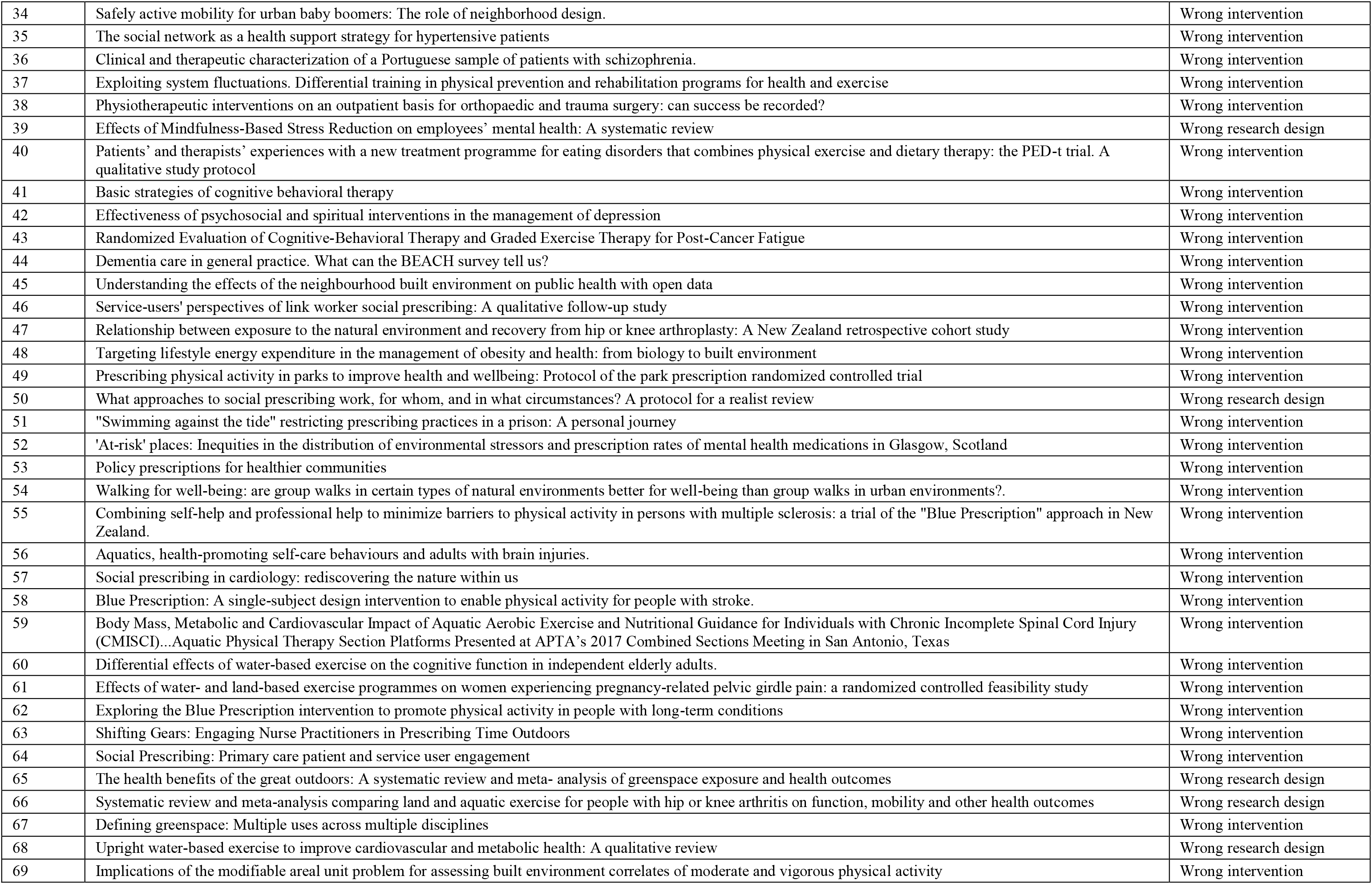
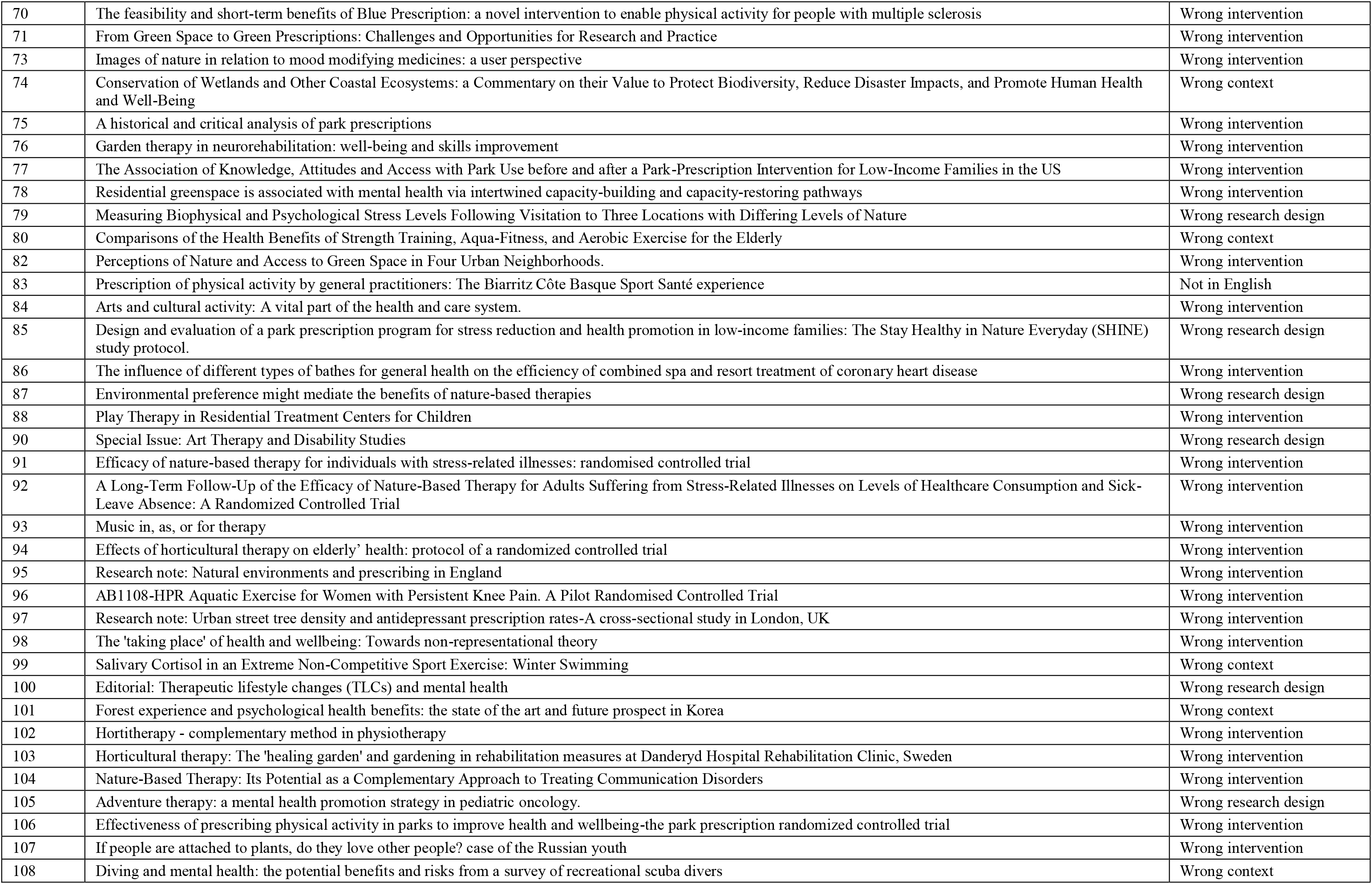
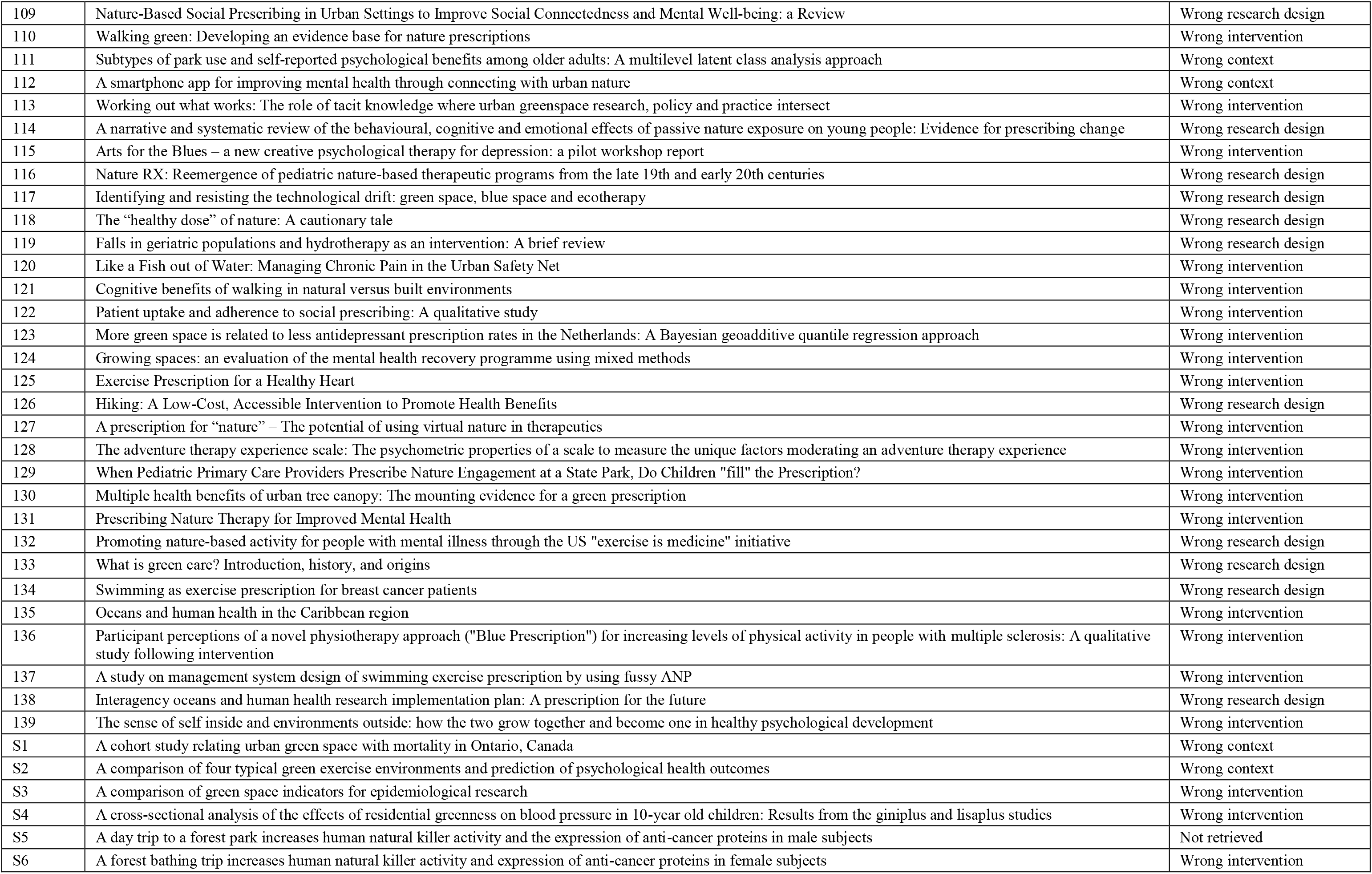
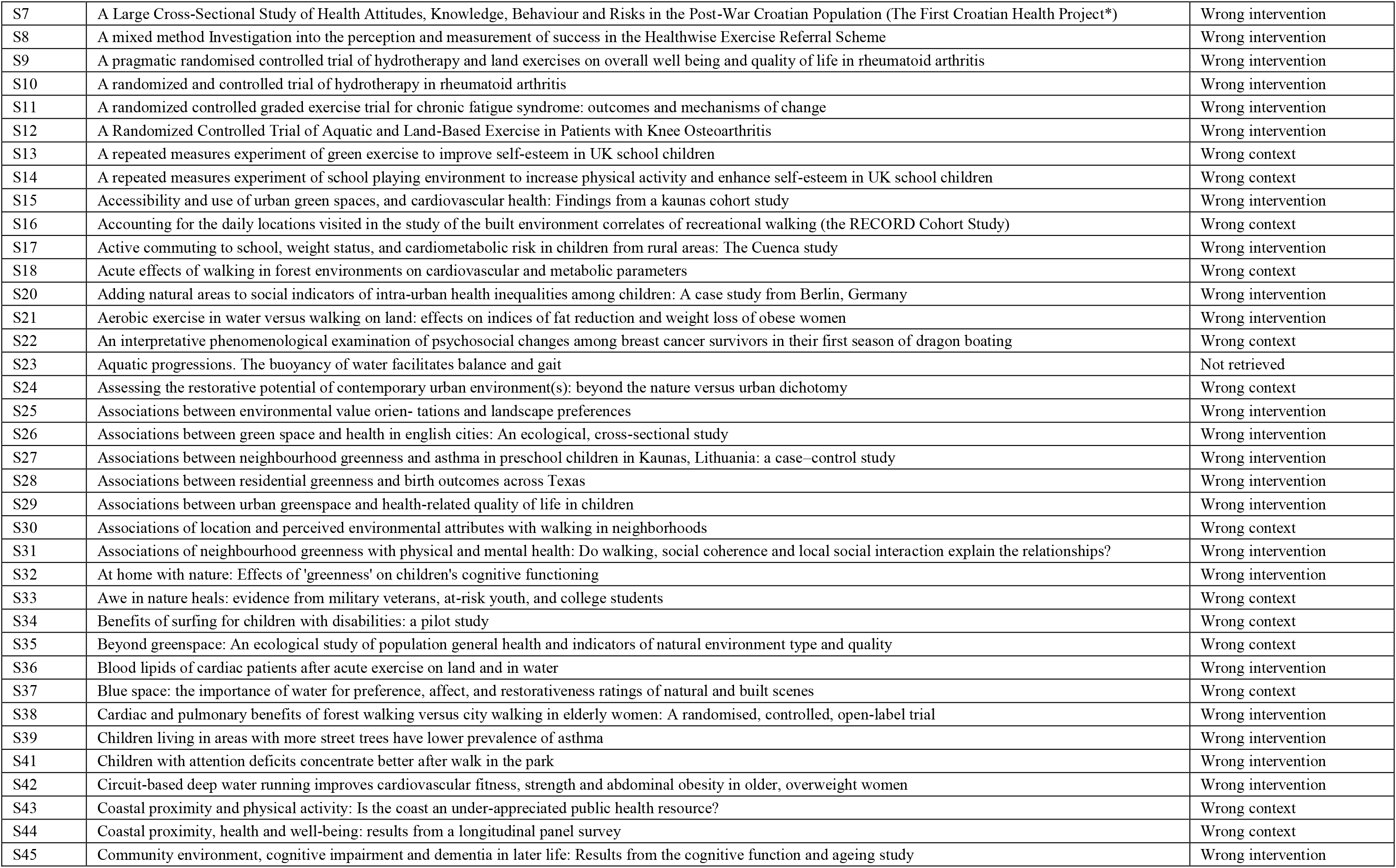
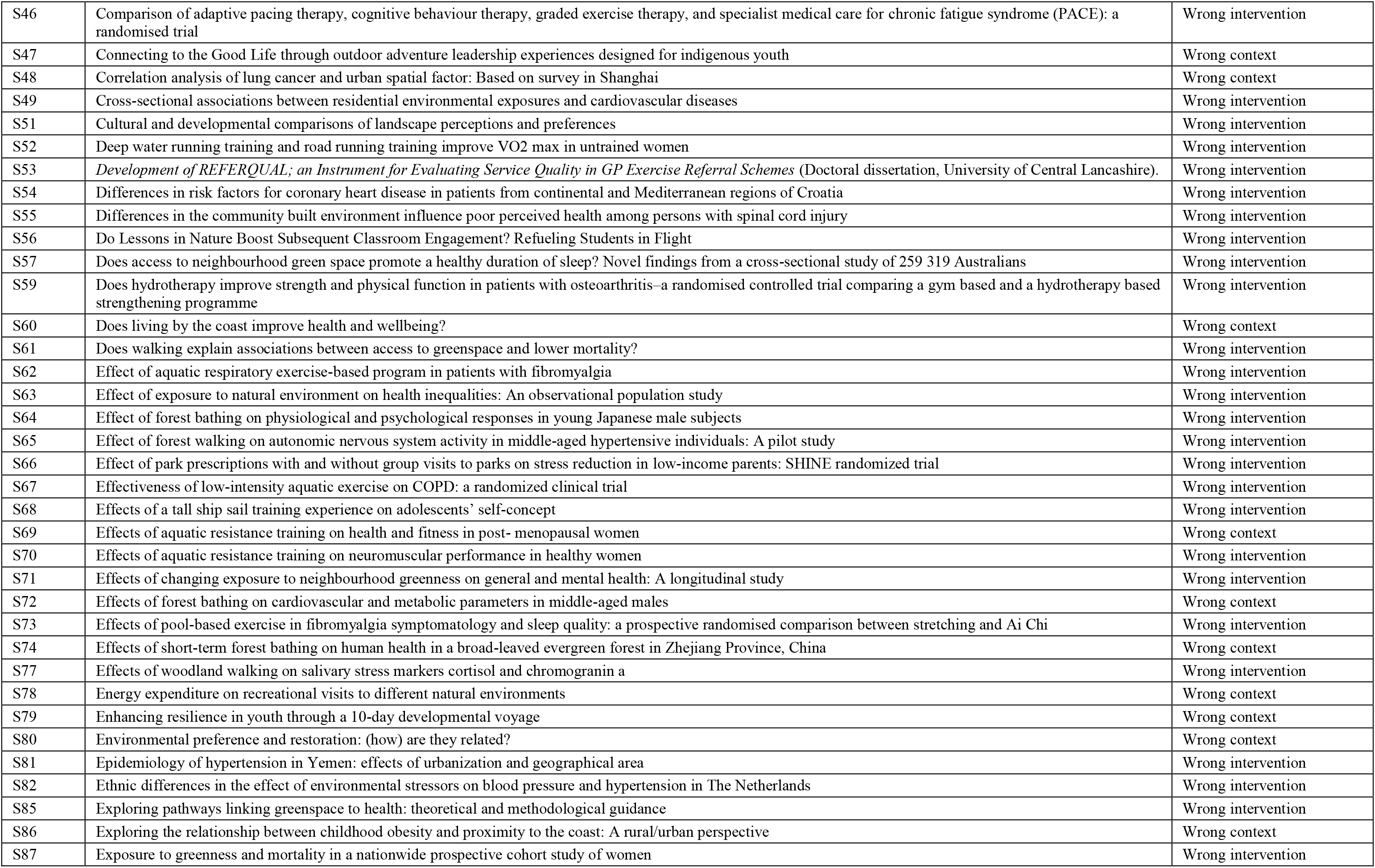
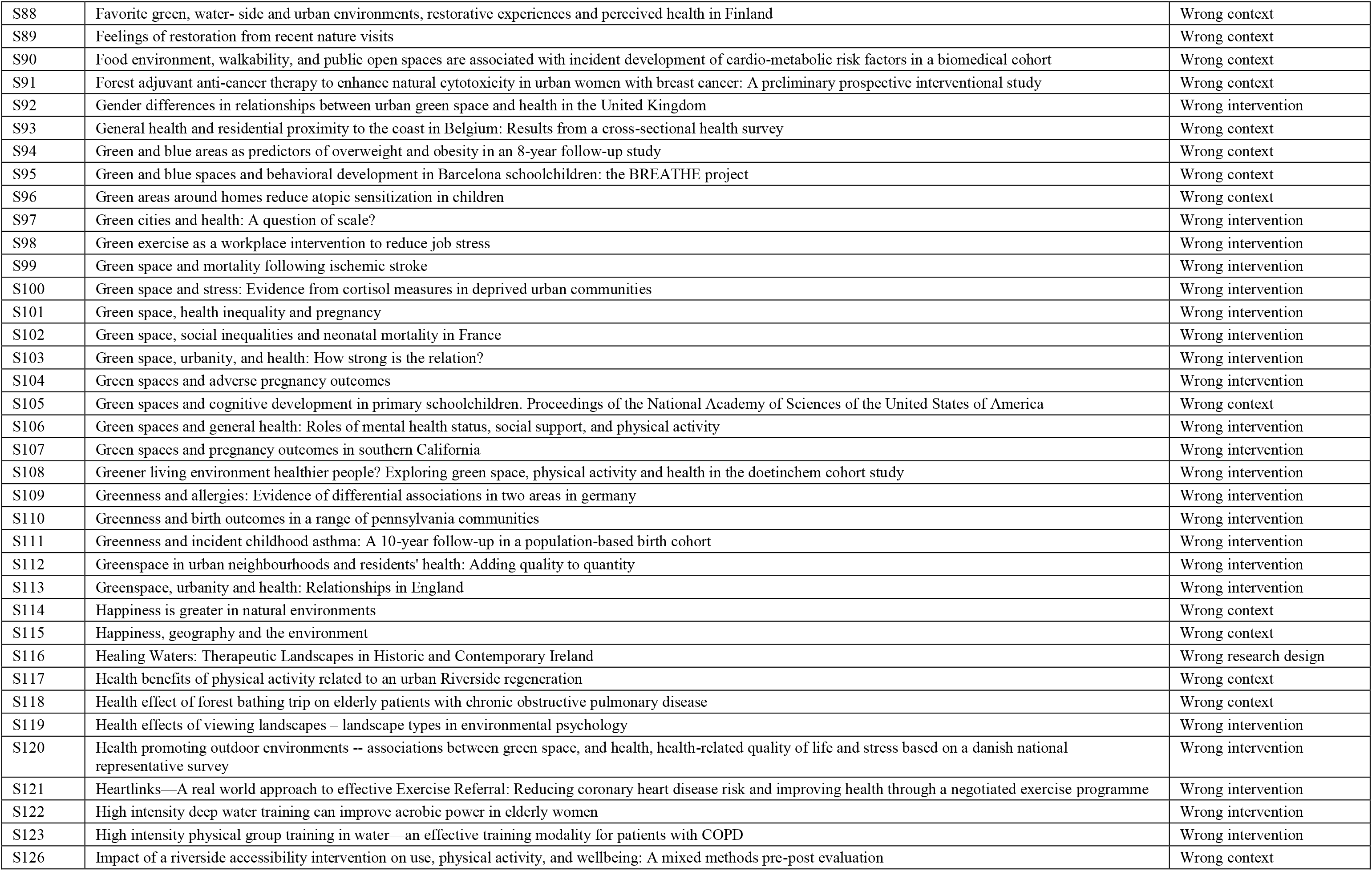
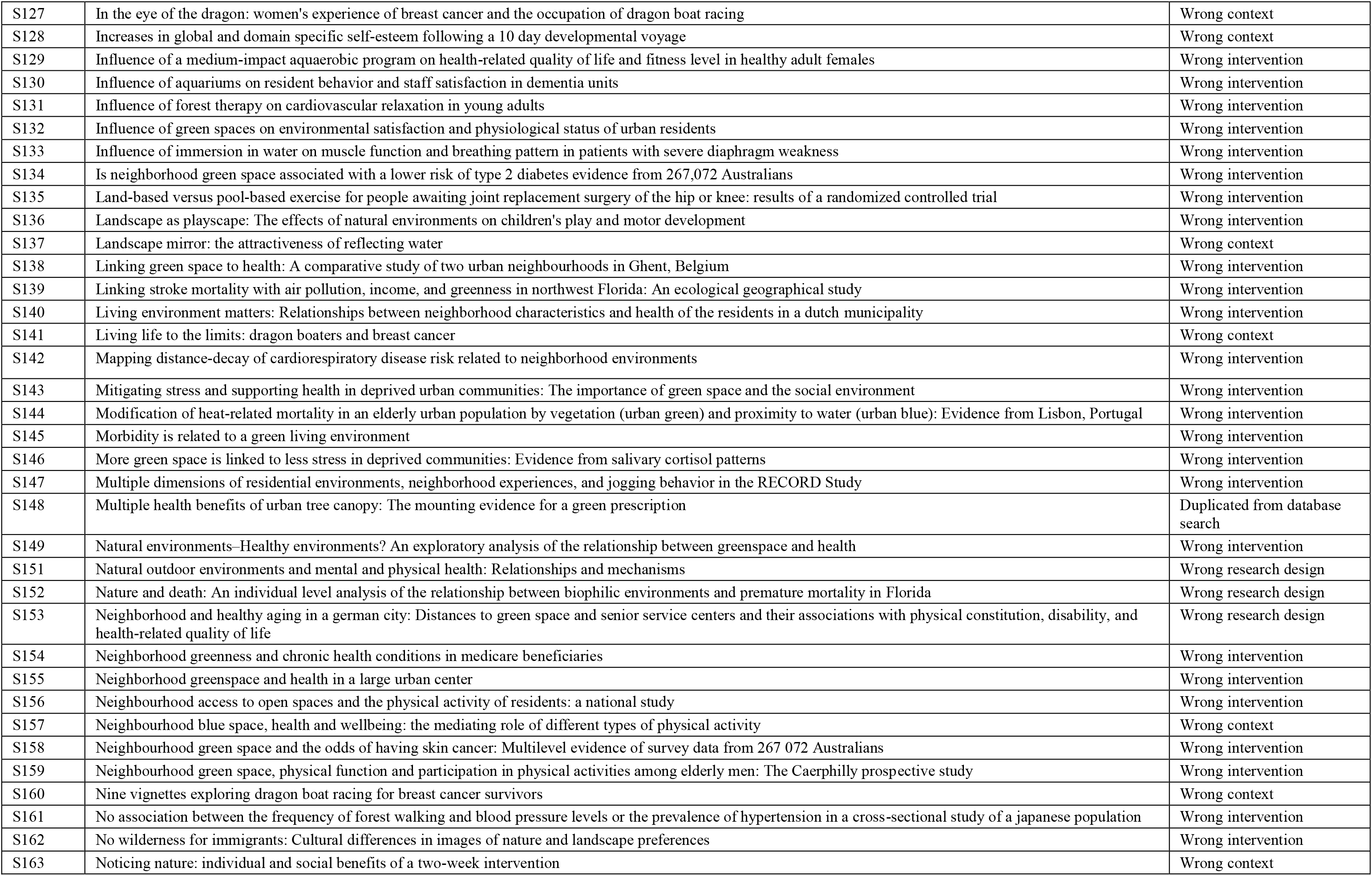
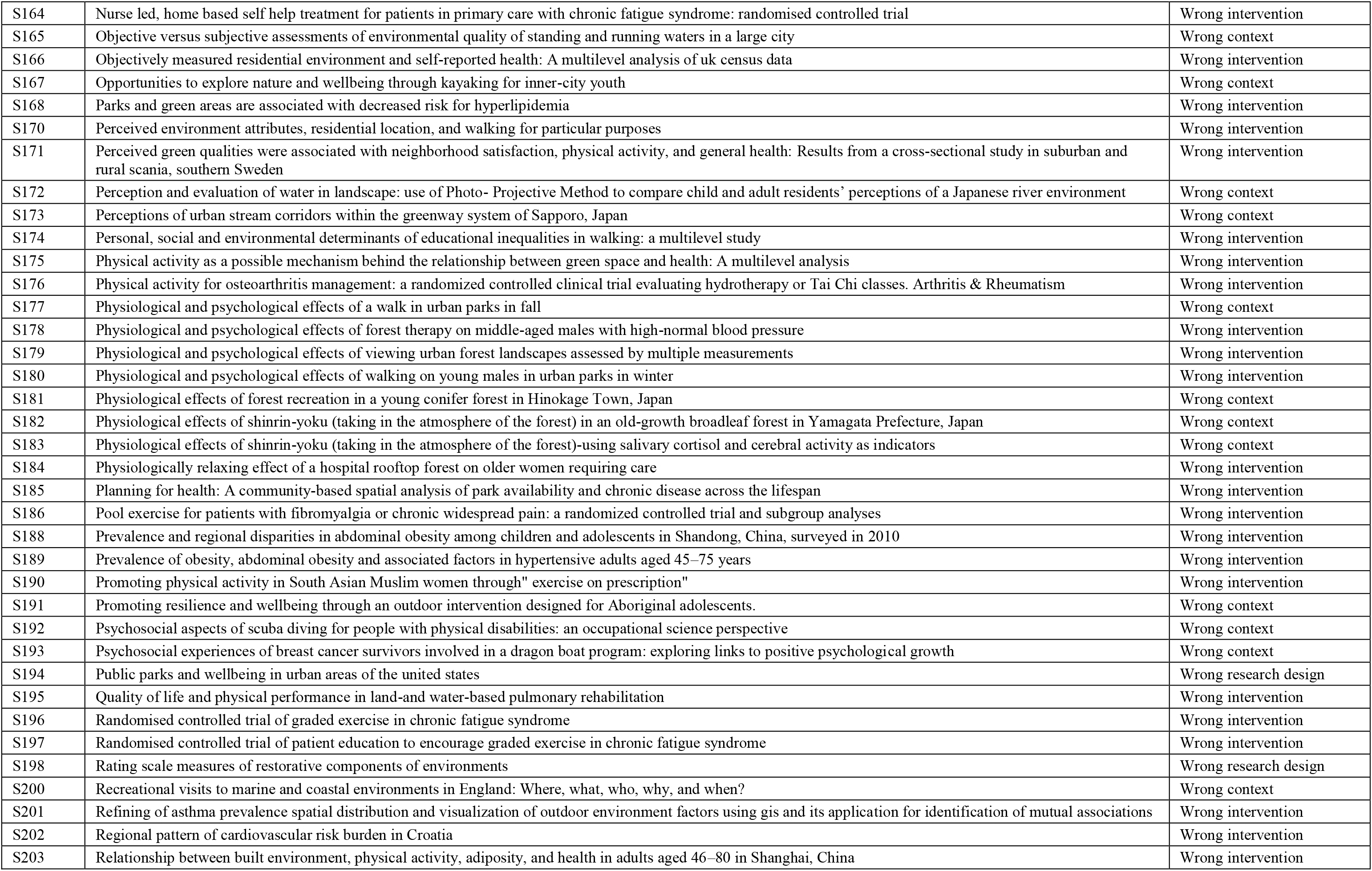
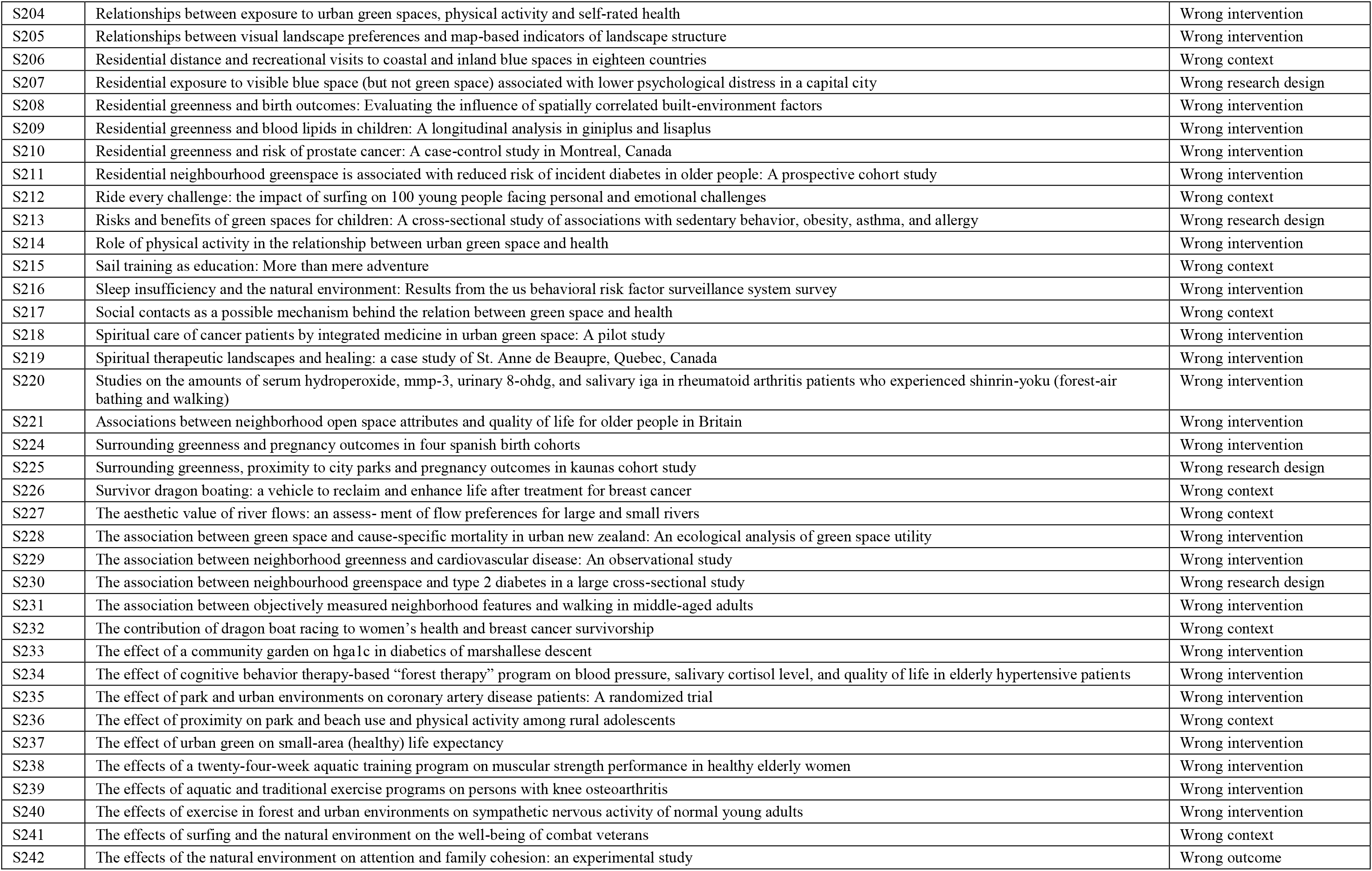
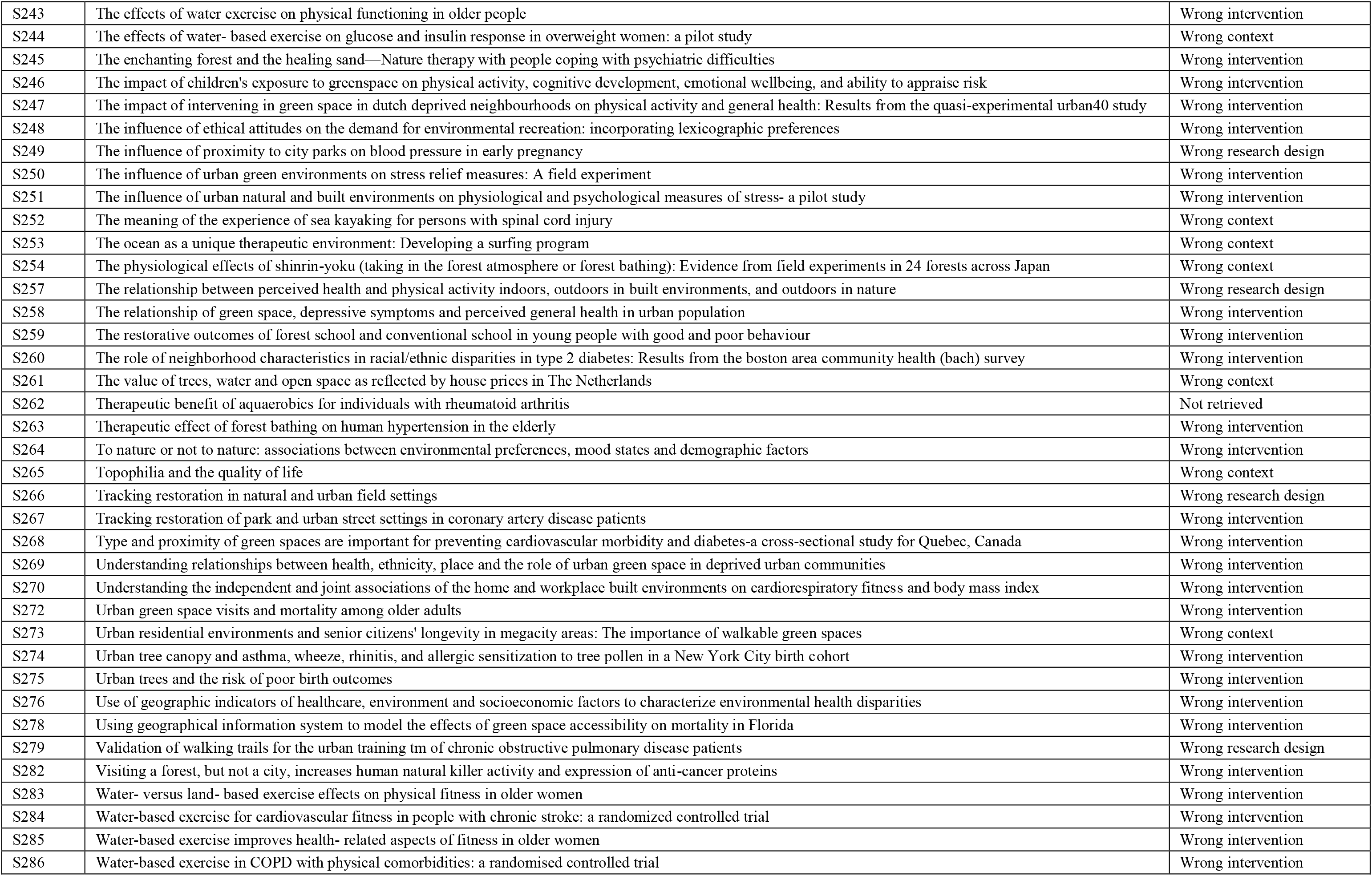
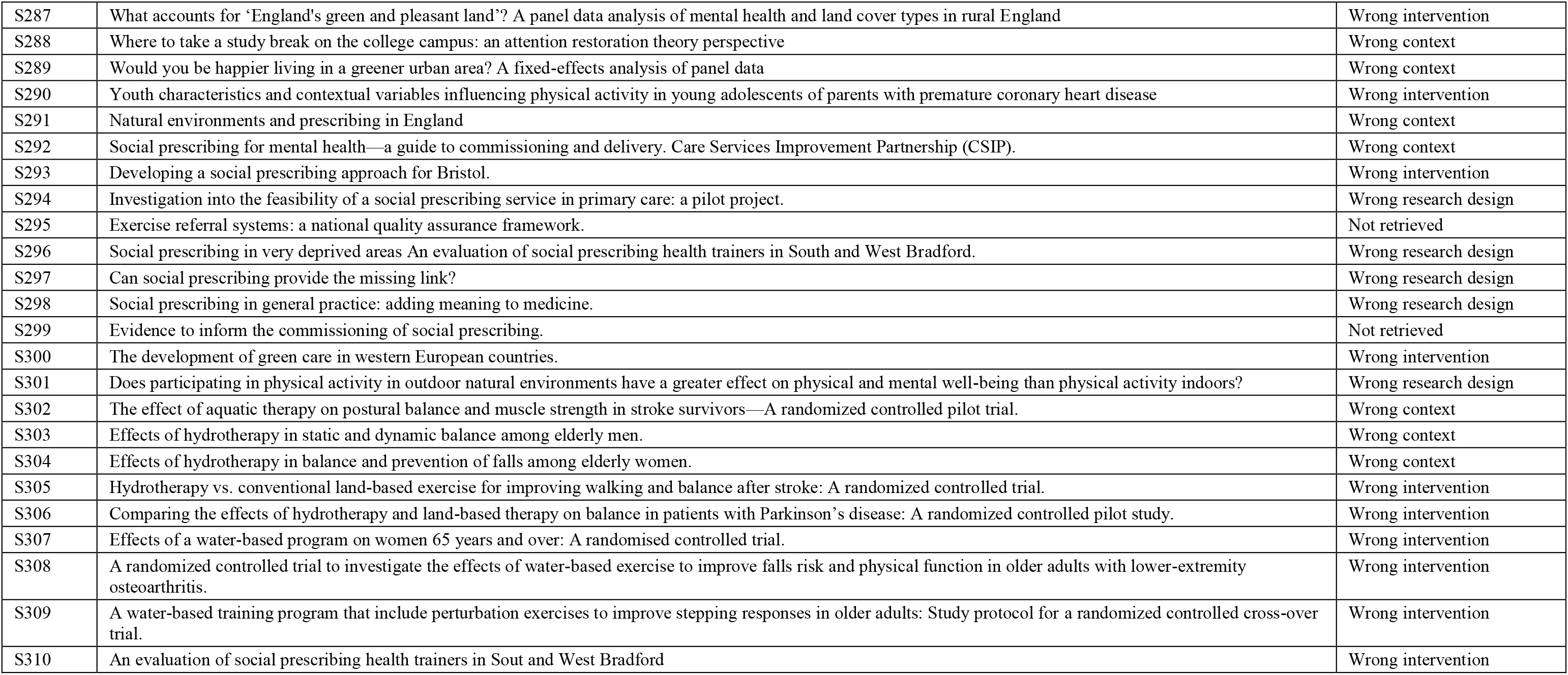
List of studies excluded at full-text screening stage with brief reasons.

## References

Agarwal R, Aggarwal AN, Gupta D. Efficacy and safety of conventional transbronchial needle aspiration in sarcoidosis: a systematic review and meta-analysis. Respiratory Care 2013; 58(4): 683–93. DOI 10.4187/respcare.02101 (accessed Aug 7, 2021)

Alejandre JC, Lynch M. “Kids Get in Shape with Nature”: A Systematic Review Exploring the Impact of Green Spaces on Childhood Obesity. J Nutr Sci Vitaminol 2020; 66(Supplement): S129–33. DOI: 10.3177/jnsv.66.S129 (accessed Mar 31, 2021)

Amoly E, Dadvand P, Forns J, et al. Green and blue spaces and behavioral development in Barcelona schoolchildren: the BREATHE project. Environ Health Perspect 2014; 122(12): 1351–8. DOI: 10.1289/ehp.1408215 (accessed Mar 30, 2021)

Astell-Burt T, Feng X. Time for ‘Green’during COVID-19? Inequities in Green and Blue Space Access, Visitation and Felt Benefits. Int J Environ Res Public Health 2021; 18(5): 2757. DOI: 10.3390/ijerph18052757 (accessed Mar 21, 2021)

Bennett JL, Van Puymbroeck M, Piatt JA, Rydell RJ. Veterans’ perceptions of benefits and important program components of a therapeutic fly-fishing program. Ther Recreation J 2014; 48(2): 169. https://www.proquest.com/docview/1553177176?pq-origsite=gscholar&fromopenview=true (accessed Aug 1, 2020)

Benton JS, Cotterill S, Anderson J, et al. A natural experimental study of improvements along an urban canal: impact on canal usage, physical activity and other wellbeing behaviours. Int J Behav Nutr Phys Act 2021; 18(1): 1–6. DOI: 10.1186/s12966-021-01088-w (accessed Aug 1, 2021)

Berg RC, Nanavati J. Realist review: current practice and future prospects. J Res Prac 2016; 12(1): R1–R1. http://jrp.icaap.org/index.php/jrp/article/view/538/449

Bertotti M, Frostick C, Hutt P, Sohanpal R, Carnes D. A realist evaluation of social prescribing: an exploration into the context and mechanisms underpinning a pathway linking primary care with the voluntary sector. Prim Health Care Res Dev 2018; 19(3): 232–45. DOI: 10.1017/S1463423617000706 (accessed Apr 21, 2021)

Bickerdike L, Booth A, Wilson PM, Farley K, Wright K. Social prescribing: less rhetoric and more reality. A systematic review of the evidence. BMJ Open 2017; 7(4). DOI: 10/1136/bmjopen-2016-013384 (accessed Apr 21, 2021)

Bratman GN, Anderson CB, Berman MG, Cochran B, de Vries S, Flanders J, et al. Nature and mental health: An ecosystem service perspective. Sci Adv 2019; 5(7): eaax0903. DOI: 10.1126/sciadv.aax0903 (accessed Mar 21, 2021)

Britten N, Stevenson FA, Barry CA, Barber N, Bradley CP. Misunderstandings in prescribing decisions in general practice: qualitative study. BMJ 2000; 320(7233): 484–8. DOI: 10.1136/bmj.320.7233.484 (accessed Mar 23, 2021)

Britton E, Kindermann G, Domegan C, Carlin C. Blue care: a systematic review of blue space interventions for health and wellbeing. Health Promot Int 2020; 35(1): 50–69. DOI: 10.1093/heapro/day103 (accessed Apr 21, 2021)

Buckley RC, Brough P. Nature, eco, and adventure therapies for mental health and chronic disease. Front Public Health 2017; 5: 220. DOI: 10.3389/fpubh.2017.00220 (accessed Apr 21, 2021)

Burgin S, Hardiman N. The direct physical, chemical and biotic impacts on Australian coastal waters due to recreational boating. Biodivers Conserv 2011; 20(4): 683–701. DOI: 10.1007/s10531-011-0003-6 (accessed Mar 21, 2021)

Cavanaugh LK, Rademacher SB. How a SURFing Social Skills Curriculum Can Impact Children with Autism Spectrum Disorders. Int J Spec Educ 2014; 15(1).

Chatterjee HJ, Camic PM, Lockyer B, Thomson LJ. Non-clinical community interventions: a systematised review of social prescribing schemes. Arts Health 2018; 10(2): 97–123. DOI: 10.1080/17533015.2017.1334002 (accessed Apr 21, 2021)

Cockburn J, Pit S. Prescribing behaviour in clinical practice: patients’ expectations and doctors’ perceptions of patients’ expectations—a questionnaire study. BMJ 1997; 315(7107): 520–3. DOI: 10.1136/bmj.315.7107.520 (accessed Mar 22, 2021)

Corley J, Okely JA, Taylor AM, et al. Home garden use during COVID-19: Associations with physical and mental wellbeing in older adults. J Environ Psychol 2021; 73:101545. DOI: 10.1016/j.jenvp.2020.101545 (accessed May 1, 2021)

Davenport J, Davenport JL. The impact of tourism and personal leisure transport on coastal environments: A review. Estuar Coast Shelf Sci 2006; 67(1–2): 280–92. DOI: 10.1016/j.ecss.2005.11.026 (accessed Mar 21, 2021)

de Matos MG, Santos AC, Fauvelet C, et al. Surfing for social integration: mental health and well-being promotion through surf therapy among institutionalized young people. HSOA J Community Med Public Health Care 2017; 4(1): 1–6. DOI: 10.24966/CMPH-1978/100026 (accessed Aug 1, 2020)

Dempsey S, Devine MT, Gillespie T, Lyons S, Nolan A. Coastal blue space and depression in older adults. Health Place 2018; 54: 110–7. DOI: 10.1016/j.healthplace.2018.09.002 (accessed Mar 23, 2021)

Dempster NR, Wildman BG, Duby J. Perception of primary care pediatricians of effectiveness, acceptability, and availability of mental health services. J Child Health Care 2015; 19(2): 195–205. DOI: 10.1177/1367493513503585 (accessed Mar 21, 2021)

Depledge MH, Bird WJ. The Blue Gym: health and wellbeing from our coasts. Mar Pollut Bull 2009; 58(7): 947–8. DOI: 10.1016/j.marpolbul.2009.04.019 (accessed Apr 21, 2021)

Dimmock K. Finding comfort in adventure: Experiences of recreational SCUBA divers. Leis Stud 2009; 28(3): 279–95. DOI: 10.1080/02614360902951674 (accessed Mar 30, 2021)

Dustin D, Bricker N, Arave J, Wall W, Wendt G. The promise of river running as a therapeutic medium for veterans coping with post-traumatic stress disorder. Ther Recreation J 2011; 45(4): 326. https://bctra.org/wp-content/uploads/tr_journals/2439-9558-1-PB.pdf (accessed Aug 1, 2021)

Elliott LR, White MP, Grellier J, Rees SE, Waters RD, Fleming LE. Recreational visits to marine and coastal environments in England: Where, what, who, why, and when? Mar Policy 2018; 97: 305–14. DOI: 10.1016/j.marpol.2018.03.013 (accessed Mar 23, 2021)

Fereday J, Muir-Cochrane E. Demonstrating rigor using thematic analysis: A hybrid approach of inductive and deductive coding and theme development. Int J Qual Methods 2006; 5(1): 80–92. DOI: 10.1177/160940690600500107 (accessed Apr 21, 2021)

Finlay J, Franke T, McKay H, Sims-Gould J. Therapeutic landscapes and wellbeing in later life: Impacts of blue and green spaces for older adults. Health Place 2015; 34: 97–106. DOI: 10.1016/j.healthplace.2015.05.001 (accessed Apr 21, 2021)

Fleischmann D, Michalewicz B, Stedje-Larsen E, et al. Surf medicine: Surfing as a means of therapy for combat-related polytrauma. J Prosthet Orthot 2011; 23(1): 27–9. DOI: 10.1097/JPO.0b013e3182065316 (accessed Aug 1, 2020)

Garrett JK, White MP, Huang J, et al. Urban blue space and health and wellbeing in Hong Kong: Results from a survey of older adults. Health Place 2019; 55: 100–10. DOI: 10.1016/j.healthplace.2018.11.003 (accessed Mar 21, 2021)

Gascon M, Zijlema W, Vert C, White MP, Nieuwenhuijsen MJ. Outdoor blue spaces, human health and well-being: A systematic review of quantitative studies. Int J Hyg Environ Health 2017; 220(8): 1207–21. DOI: 10.1016/j/ijheh.2017.08.004 (accessed Apr 21, 2021)

Geng DC, Innes J, Wu W, Wang G. Impacts of COVID-19 pandemic on urban park visitation: a global analysis. J For Res 2021; 32(2): 553–67. DOI: 10.1007/s11676-020-01249-w (accessed Mar 23, 2021)

Georgiou M, Morison G, Smith N, Tieges Z, Chastin S. Mechanisms of Impact of Blue Spaces on Human Health: A Systematic Literature Review and Meta-Analysis. Int J Environ Res Public Health 2021; 18(5): 2486. DOI: 10.3390/ijerph18052486 (accessed Jul 7, 2021)

Godfrey C, Devine-Wright H, Taylor J. The positive impact of structured surfing courses on the wellbeing of vulnerable young people. Community Pract 2015; 88(1): 26–9. https://pubmed.ncbi.nlm.nih.gov/26357740/ (accessed Aug 1, 2020)

Graham AL, Cooke SJ. The effects of noise disturbance from various recreational boating activities common to inland waters on the cardiac physiology of a freshwater fish, the largemouth bass (Micropterus salmoides). Aquat Conserv 2008; 18(7): 1315–24. DOI: 10.1002/aqc.941 (accessed Mar 21, 2021)

Ha JF, Longnecker N. Doctor-patient communication: a review. Ochsner J 2010; 10(1): 38–43. www.ochsnerjournal.org/content/ochjnl/10/1/38.full.pdf (accessed Mar 21, 2021)

Hartig T, Mitchell R, de Vries S, Frumkin H. Nature and Health. Annu Rev Public Health 2014; 35: 207–28. DOI: 10.1146/annurev-pubhealth-032013-182443 (accessed Aug 10, 2021)

Helbich M, Yao Y, Liu Y, Zhang J, Liu P, Wang R. Using deep learning to examine street view green and blue spaces and their associations with geriatric depression in Beijing, China. Environ Int 2019; 126: 107–17. DOI: 10.1016/j.envint.2019.02.013 (accessed Mar 21, 2021)

Heo S, Lim CC, Bell ML. Relationships between Local Green Space and Human Mobility Patterns during COVID-19 for Maryland and California, USA. Sustainability 2020; 12(22): 9401. DOI: 10.3390/su12229401 (accessed Mar 21, 2021)

Hignett A, White MP, Pahl S, Jenkin R, Froy ML. Evaluation of a surfing programme designed to increase personal well-being and connectedness to the natural environment among ‘at risk’ young people. J Adventure Educ Outdoor Lear 2017; 18(1): 53–69. DOI: 10.1080/14729679.2017.1326829 (accessed Aug 1, 2020)

Hubbard G, Daas CD, Johnston M, Murchie P, Thompson CW, Dixon D. Are Rurality, Area Deprivation, Access to Outside Space, and Green Space Associated with Mental Health during the COVID-19 Pandemic? A Cross Sectional Study (CHARIS-E). Int J Environ Res Public Health 2021; 18(8): 3869. DOI: 10.3390/ijerph18083869 (accessed May 1, 2021)

Husk K, Blockley K, Lovell R, et al. What approaches to social prescribing work, for whom, and in what circumstances? A realist review. Health Soc Care Community 2020; 28(2): 309–24. DOI: 10.1111/hsc.12839 (accessed Apr 7, 2021)

Huttunen P, Kokko L, Ylijukuri V. Winter swimming improves general well-being. Int J Circumpolar Health 2004; 63(2): 140–4. DOI: 10.3402/ijch.v63i2.17700 (accessed Mar 30, 2021)

Jalali S, Wohlin C. Systematic literature studies: database searches vs. backward snowballing. In: Proceedings of the 2012 ACM-IEEE International Symposium on Empirical Software Engineering and Measurement. 2012. pp.29–38.

James AK, Hess P, Perkins ME, Taveras EM, Scirica CS. Prescribing outdoor play: outdoors Rx. Clin Pediatr 2017; 56(6): 519–24. DOI: 10.1177/0009922816677805 (accessed Aug 1, 2020)

Keniger LE, Gaston KJ, Irvine KN, Fuller RA. What are the benefits of interacting with nature? Int J Environ Res Public Health 2013; 10(3): 913–935. DOI: 10.3390/ijerph10030913 (accessed Aug 10, 2021)

Kmet LM, Cook LS, Lee RC. Standard quality assessment criteria for evaluating primary research papers from a variety of fields. Alberta Heritage Foundation for Medical Research. HTA Initiative #13. 2004.

Knapp M, McDaid D, Parsonage M. Mental Health Promotion and Mental Illness Prevention: The Economic Case. Department of Health London. 2011. 61

Leavell MA, Leiferman JA, Gascon M, Braddick F, Gonzalez JC, Litt JS. Nature-based social prescribing in urban settings to improve social connectedness and mental well-being: a review. Curr Environ Health Rep 2019; 6(4): 297–308. DOI: 10.1007/s40572-019-00251-7 (accessed Apr 21, 2021)

Lee L, Packer TL, Tang SH, Girdler S. Self-management education programs for age-related macular degeneration: A systematic review. Australas J Ageing 2008; 27(4): 170–6. DOI: 10.1111/j.1741-6612-2008.00298.x (accessed Apr 21, 2021)

Leeworthy VR. Preliminary Estimates from Version 1-6: Coastal Recreation Participation. US Department of Commerce. 2001.

Little P, Everitt H, Williamson I, et al. Preferences of patients for patient centred approach to consultation in primary care: observational study. BMJ 2001; 322(7284): 468. DOI: 10.1136/bmj.322.7284.468 (accessed Mar 22, 2021)

Lopes JT. Adapted surfing as a tool to promote inclusion and rising disability awareness in Portugal. J Sport Dev 2015; 3(5): 4–10.https://jsfd.wordpress.com/2020/08/lopes.adapated.surfing.inclusion.pdf (accessed Aug 1, 2020)

Mackay GJ, Neill JT. The effect of “green exercise” on state anxiety and the role of exercise duration, intensity, and greenness: A quasi-experimental study. Psychol Sport Exerc 2010; 11(3): 238–45. DOI: 10.1016/j.psychsport.2010.01.002 (accessed Mar 29, 2021)

Maharaj S, Harding R. The needs, models of care, interventions and outcomes of palliative care in the Caribbean: a systematic review of the evidence. BMC Palliat Care 2016; 15(1): 1–20. DOI: 10.1186/s12904-016-0079-6 (accessed Apr 21, 2021)

Markevych I, Schoierer J, Hartig T, et al. Exploring pathways linking greenspace to health: theoretical and methodological guidance. Environ Res 2017; 158: 301–317. DOI: 10.1016/j.envres.2017.06.028 (accessed Aug 10, 2021)

Masterton W, Carver H, Parkes T, Park K. Greenspace interventions for mental health in clinical and non-clinical populations: What works, for whom, and in what circumstances? Health Place 2020; 64(102338): 102338. DOI: 10.1016/j/healthplace.2020.102338 (accessed Aug 7, 2021)

Maund PR, Irvine KN, Reeves J, et al. Wetlands for wellbeing: piloting a nature-based health intervention for the management of anxiety and depression. Int J Environ Res Public Health 2019; 16(22): 4413. DOI: 10.3390/ijerph16224413 (accessed Aug 1, 2020)

Maxwell RJ, Zolderdo AJ, de Bruijn R, et al. Does motor noise from recreational boats alter parental care behaviour of a nesting freshwater fish? Aquatic Conserv: Mar Freshw Ecosyst 2018; 28: 969–978. DOI: 10.1002/aqc.2915 (accessed Mar 21, 2021)

McHale S, Pearsons A, Neubeck L, Hanson CL. Green Health Partnerships in Scotland; Pathways for social prescribing and physical activity referral. Int J Environ Res Public Health 2020; 17(18). DOI: 10.3390/ijerph17186832 (accessed Mar 21, 2021)

Michie S, van Stralen MM, West R. The behaviour change wheel: a new method for characterising and designing behaviour change interventions. Implement Sci 2011; 6(1): 42. http://www.implementationsicne.com/content/6/1/42 (accessed Apr 21, 2021)

Mowatt RA, Bennett J. Veteran stories, PTSD effects and therapeutic fly-fishing. Ther Recreation J 2011; 45: 286–308.

NHS England. The NHS long term plan. NHS England. 2019. https://www.longtermplan.nhs.uk/wp-content/uploads/2019/08/nhs-long-term-plan-version-1.2.pdf (accessed Mar 21, 2021)

Ofiara DD, Brown B. Assessment of economic losses to recreational activities from 1988 marine pollution events and assessment of economic losses from long-term contamination of fish within New York bight to New Jersey. Mar Pollut Bull 1999; 20(4): 990–1004. DOI: 10.1016/S0025-326X(99)00123-X (accessed Mar 21, 2021)

Ouzzani M, Hammady H, Fedorowicz Z, Elmagarmid A. Rayyan—a web and mobile app for systematic reviews. Syst Rev 2016; 5(1): 1–0. DOI: 10.1186/s13643-016-0384-4 (accessed Apr 21, 2021)

Page MJ, McKenzie JE, Bossuyt PM, et al. The PRISMA 2020 statement: an updated guideline for reporting systematic reviews. BMJ 2021; 372. DOI: 10.1136/bmj.n71 (accessed Aug 7, 2021)

Pawson R, Bellamy JL. Realist synthesis: an explanatory focus for systematic review. In: Popay J, editor. Moving beyond effectiveness in evidence synthesis. London: National Institute for Health and Clinical Excellence; 2006. p. 83–93.

Pawson R, Greenhalgh T, Harvey G, Walshe K. Realist review-a new method of systematic review designed for complex policy interventions. J Health Serv Res Pol 2005; 10(1_suppl): 21–34. DOI: 10.1258/1355819054308530 (accessed Jul 7, 2021)

Pescheny JV, Pappas Y, Randhawa G. Facilitators and barriers of implementing and delivering social prescribing services: a systematic review. BMC Health Ser Res 2018; 18(1): 1–4. DOI: 10.1186/s12913-018-2893-4 (accessed Apr 21, 2021)

Pilkington K, Loef M, Polley M. Searching for real-world effectiveness of health care innovations: scoping study of social prescribing for diabetes. J Med Internet Res 2017; 19(2): e20. DOI: 10.2196/jmir.6431 (accessed Apr 21, 2021)

Pouso S, Borja Á, Fleming LE, Gómez-Baggethun E, White MP, Uyarra MC. Contact with blue-green spaces during the COVID-19 pandemic lockdown beneficial for mental health. Sci Total Environ 2021; 756:143984. DOI: 10.1016/j.scitotenv.2020.143984 (accessed Jul 7, 2021)

Pretty J, Peacock J, Hine R, Sellens M, South N, Griffin M. Green exercise in the UK countryside: Effects on health and psychological well-being, and implications for policy and planning. J Environ Plan Manag 2007; 50(2): 211–31. DOI: 10.1080/09640560601156466 (accessed Mar 29, 2021)

Razai MS, Oakeshott P, Kankam H, Galea S, Stokes-Lampard H. Mitigating the psychological effects of social isolation during the COVID-19 pandemic. BMJ 2020; 369. DOI: 10.1136/bmj.m1904 (accessed Mar 21, 2021)

Razani N, Morshed S, Kohn MA, et al. Effect of park prescriptions with and without group visits to parks on stress reduction in low-income parents: SHINE randomized trial. PloS One 2018; 13(2): e0192921. DOI: 10.1371/journal/pone.0192921 (accessed Aug 1, 2020)

Roberts L, Jones G, Brooks R. Why Do You Ride?: A Characterization of Mountain Bikers, Their Engagement Methods, and Perceived Links to Mental Health and Well-Being. Front Psychol 2018; 9: 1642. DOI: 10.3389/fpsyg.2018.01642 (accessed Mar 30, 2021)

Rogers CM, Mallinson T, Peppers D. High-intensity sports for posttraumatic stress disorder and depression: Feasibility study of ocean therapy with veterans of Operation Enduring Freedom and Operation Iraqi Freedom. Am J of Occup Ther 2014; 68(4): 395–404. DOI:10.5014/ajot/2014.011221 (accessed Aug 1, 2020)

Rycroft-Malone J, McCormack B, Hutchinson AM, DeCorby K, Bucknall TK, Kent B, et al. Realist synthesis: illustrating the method for implementation research. Implement Sci 2012; 7(1): 33. https://www.implementationscience.com/content/7/1/33 (accessed Jul 7, 2021)

Schardt C, Adams MB, Owens T, Keitz S, Fontelo P. Utilization of the PICO framework to improve searching PubMed for clinical questions. BMC Med Inform Decis Mak 2007; 7(1): 1–6. DOI: 10.1186/1472-6947-7-16 (accessed Apr 21, 2021)

Sharma M, Largo-Wight E, Kanekar A, Kusumoto H, Hooper S, Nahar VK. Using the multi-theory model (MTM) of health behavior change to explain intentional outdoor nature contact behavior among college students. Int J Environ Res Public Health 2020; 17(17): 6104. DOI: 10.3390/pharmacy9030126 (accessed Apr 21, 2021)

Tester-Jones M, White MP, Elliott LR, et al. Results from an 18 country cross-sectional study examining experiences of nature for people with common mental health disorders. Sci Rep 2020; 10(1): 1–1. DOI: 10.1038/s41598-020-75825-9 (accessed Mar 31, 2021)

Teuton, J. Social prescribing for mental health: background paper. NHS Health Scotland. 2015. www.healthscotland.scot/media/2067/social-prescribing-for-mental-health-background-paper.pdf (accessed Apr 21, 2021)

Thomas J, Harden A. Methods for the thematic synthesis of qualitative research in systematic reviews. BMC Med Res Methodol 2008; 8(1): 1–0. DOI: 10.1186/1471-2288-8-45 (accessed Apr 21, 2021)

Tieges Z, McGregor D, Georgiou M, et al. The Impact of Regeneration and Climate Adaptations of Urban Green–Blue Assets on All-Cause Mortality: A 17-Year Longitudinal Study. Int J Environ Res Public Health 2020; 17(12): 4577. DOI: 10.3390/ijerph17124577 (accessed Jul 7, 2021)

United Nations’ Food and Agriculture Organisation, European Union, Organisation for Economic Co-operation and Development, World Bank Group. System of Environmental-Economic Accounting 2012–Experimental Ecosystem Accounting. United Nations. 2014. https://seea.un.org/sites/seea.un.org/files/seea_eea_final_en_1.pdf (accessed Aug 10, 2021)

Van den Berg M,., W. endel-Vos W-VW,, Van Poppel M., H. K, Mechelen W., J M et al. Health benefits of green spaces in the living environment: a systematic review of epidemiological studies. Urban For Urban Green 2015; 14: 806–816. DOI: 10.1016/j.ufug.2015.07.008 (accessed Mar 21, 2021)

van Tulleken C, Tipton M, Massey H, Harper CM. Open water swimming as a treatment for major depressive disorder. Case Reports 2018; 2018: bcr-2018. DOI: 10.1136/bcr-2018-225007 (accessed Mar 30, 2021)

Varkey B. Principles of clinical ethics and their application to practice. Med Princ Pract 2021; 30(1): 17–28. DOI 10.1159/000509119 (accessed Mar 9, 2021)

Vella EJ, Milligan B, Bennett JL. Participation in outdoor recreation program predicts improved psychosocial well-being among veterans with post-traumatic stress disorder: A pilot study. Mil Med 2013; 178(3): 254–60. DOI: 10.7205/MILMED-D-12-00308 (accessed Aug 1, 2020)

Vert C, Carrasco-Turigas G, Zijlema W, et al. Impact of a riverside accessibility intervention on use, physical activity, and wellbeing: A mixed methods pre-post evaluation. Landsc Urban Plan 2019; 190: 103611. DOI: 10.1016/j.landurbplan.2019.103611 (accessed Mar 27, 2021)

Völker S, Kistemann T. The impact of blue space on human health and well-being - Salutogenetic health effects of inland surface waters: a review. Int J Hyg Environ Health 2011; 214(6): 449–60. DOI: 10.1016/j.ijheh.2011.05.001 (accessed Apr 21, 2021)

White MP, Alcock I, Grellier J, et al. Spending at least 120 minutes a week in nature is associated with good health and wellbeing. Sci Rep 2019; 9(1): 1–1. DOI: 10.1038/s41598-019-44097-3 (accessed Mar 22, 2021)

White MP, Elliott LR, Gascon M, Roberts B, Fleming LE. Blue space, health and well-being: A narrative overview and synthesis of potential benefits. Environ Res 2020; 22: 110169. DOI: 10.1016/j.envres.2020.110169 (accessed Apr 21, 2021)

White R, Abraham C, Smith JR, White M, Staiger PK. Recovery under sail: rehabilitation clients’ experience of a sail training voyage. Addict Res Theory 2016; 24(5): 355–65. DOI: 10.3109/16066359.2015.1123252 (accessed Aug 1, 2020)

Wolch J, Zhang J. Beach recreation, cultural diversity and attitudes toward nature. J Leis Res 2004; 36(3): 414–43. DOI: 10.1080/00222216.20024.11950030 (accessed Mar 23, 2021)

Wood SL, Demougin PR, Higgins S, Husk K, Wheeler BW, White M. Exploring the relationship between childhood obesity and proximity to the coast: A rural/urban perspective. Health Place 2016; 40: 129–36. DOI: 10.1016/j.healthplace.2016.05.010 (accessed Mar 22, 2021)

Yang Y, Wang L, Passmore H-A, Zhang J, Zhu L, Cai H. Viewing nature scenes reduces the pain of social ostracism. J Soc Psychol 2021; 161(2): 197–215. DOI: 10.1080/00224545.2020.1784826 (accessed May 1, 2021)

